# The contribution of coding variants to the heritability of multiple cancer types using UK Biobank whole-exome sequencing data

**DOI:** 10.1101/2024.09.30.24314593

**Authors:** Naomi Wilcox, Jonathan P. Tyrer, Joe Dennis, Xin Yang, John R. B. Perry, Eugene J. Gardner, Douglas F. Easton

**Affiliations:** Centre for Cancer Genetic Epidemiology, Department of Public Health and Primary Care. University of Cambridge. UK; Metabolic Research Laboratory, Wellcome-MRC Institute of Metabolic Science. University of Cambridge. UK; MRC Epidemiology Unit, Wellcome-MRC Institute of Metabolic Science. University of Cambridge. UK; Centre for Cancer Genetic Epidemiology, Department of Oncology. University of Cambridge. UK

## Abstract

Genome-wide association studies have been highly successful at identifying common variants associated with cancer, however, they do not explain all the inherited risk of cancer. Family-based studies, targeted sequencing and, more recently, exome-wide association studies have identified rare coding variants in some genes associated with cancer risk, but the overall contribution of these variants to the heritability of cancer is less clear. Here we describe a method to estimate the genome-wide contribution of rare coding variants to heritability that fits models to the burden effect sizes using an empirical Bayesian approach. We apply this method to the burden of protein-truncating variants in over 15,000 genes for 11 cancers in UK Biobank, using whole-exome sequencing data on over 400,000 individuals. We extend the method to consider the overlap of genes contributing to pairs of cancers. We found ovarian cancer to have the greatest proportion of heritability attributable to protein-truncating variants in genes (46%). The joint cancer models highlight significant clustering of cancer types, including a near complete overlap in susceptibility genes for breast, ovarian, prostate and pancreatic cancer. Our results provide insights into the contribution of rare coding variants to the heritability of cancer and identify additional genes with strong evidence of susceptibility to multiple cancer types.

## Introduction

Genome-wide association studies have been highly successful at identifying common variants associated with disease. Increasingly, association studies are being extended to study rare variants using next-generation sequencing methods. For example, for breast cancer, GWAS have identified over 300 common susceptibility loci^1-3^, while rare variants in *ATM, BARD1, BRCA1, BRCA2, CHEK2, RAD51C, RAD51D, PALB2* and *TP53* have been identified through linkage or targeted sequencing studies^4^. Exome-wide analysis has recently additionally identified rare variants in MAP3K1 to be associated with breast cancer risk^5^. Similarly, for bowel cancer, GWAS have identified over 200 common susceptibility loci^6^, and rare variants have been identified in mismatch repair (MMR) genes including *MSH2, MSH6, MLH1, PMS2*, as well other genes including *APC*^7^. The increasing availability of whole-exome and whole-genome sequencing data is enabling exome- and genome-wide analysis of rare variants, and the discovery of novel rare variants associated with cancer risk.

For common variants, GWAS data can be used to estimate the overall contribution to heritability, using methods including LD score regression (LDSC)^8^, which uses GWAS summary results, and TGCA^9^. LDSC has been used to estimate that common variants explain ∼41% of the familial relative risk of breast cancer and ∼73% of colorectal cancer^2,6^. This method has also been extended to estimate the genetic correlation between traits^10^, which has shown significant correlations in cancer susceptibility, for example for breast and ovarian cancer, and breast and lung cancer^11^. An analogous question is: what is the overall contribution of rare coding variants to cancer heritability? While some genes, such as *BRCA1* and *BRCA2*, have long been known to make a significant contribution to the familial aggregation of certain cancers^5,12^, the more general question has not been definitively answered since most genes have not been extensively evaluated in association studies.

We previously described a method for evaluating the contribution of the gene-wise burden of rare coding variants to cancer heritability that fits models to the burden effect sizes using an empirical Bayesian approach^5^. This approach can be implemented using gene burden summary statistics and is not computationally intensive. We previously applied this method to the burden of protein-truncating variants (PTVs) in genes and breast cancer risk using data from the UK Biobank and Breast Cancer Association Consortium (BCAC)^5^. Here we apply this method to 11 different cancer types using data from UK Biobank. We extend this method to consider the overlap of genes contributing to pairs of cancers and evaluate the correlation of coding variant heritability among cancers. This method can be used to derive posterior probabilities that each gene is cancer associated.

## Material and Methods

### Material – UK Biobank

UK Biobank is a population-based prospective cohort study of more than 500,000 individuals. More detailed information on the UK Biobank is given elsewhere^13,14^. WES data for 450,000 samples were released in October 2021 and accessed via the UK Biobank DNANexus platform^15^. QC metrics were applied to Variant Call Format (VCF) files as described by Gardner et Al, including genotype level filters for depth and genotype quality^16^. Other filters, including exclusion of samples with disagreement between genetically determined and self-reported sex or excess relatives were applied as described elsewhere^17^. The final dataset for analysis included 419,307 samples with 227,393 females and 191,914 males.

Cancer cases were determined by linkage to national cancer registration data (NCRAS) and by selecting the appropriate ICD-10 codes (Table 1). For breast cancer, we also included self-reported cancer. Both prevalent and incident cases were included. Only cancers which were an individual’s first or second diagnosed cancer were included as cases. The numbers of cases for males and females for each cancer are provided in Table 1. The 11 cancers we considered were the most common solid tumors by incident cancer diagnoses in UK Biobank, as per the UK Biobank Malignant Cancer Summary Report^18^.

**Table 1.** Cancer ICD10 codes, case and control counts, and the model used for analysis.

|  | ICD10 code(s) | Female |  | Male |  | FH variable | Model |
| --- | --- | --- | --- | --- | --- | --- | --- |
|  |  | Control | Case | Control | Case |  |  |
| Breast | C50, D05 | 209,435 | 17,958 | 191,820 | 94 | Yes | 3 |
| Lung | C34 | 225,612 | 1,781 | 189,959 | 1,955 | Yes | 3 |
| Prostate | C61 | 227,393 | 0 | 180,214 | 11,700 | Yes | 3 |
| Bowel | C18, C19, C20, C21 | 224,404 | 2,989 | 187,956 | 3,958 | Yes | 3 |
| Pancreatic | C25 | 226,900 | 493 | 191,326 | 588 | No | 2 |
| Endometrial | C54 | 225,419 | 1,974 | 191,914 | 0 | No | 1 |
| Ovarian | C56 | 226,021 | 1,372 | 191,914 | 0 | No | 1 |
| Oesophagus | C15 | 227,118 | 275 | 191,167 | 747 | No | 2 |
| Kidney | C64 | 226,774 | 619 | 190,862 | 1,052 | No | 2 |
| Bladder | C67 | 227,072 | 321 | 190,780 | 1,134 | No | 2 |
| Malignant Melanoma | C43 | 225,111 | 2,282 | 189,986 | 1,928 | No | 2 |

The Ensembl Variant Effect Predictor (VEP) was used to annotate variants, including the 1000 genomes phase 3 allele frequency, sequence ontology variant consequences and exon/intron number^19^. Annotation files were used to identify PTVs. PTVs in the last exon of each gene and the last 50 bp of the penultimate exon were excluded as these are generally predicted to escape Nonsense-Mediated mRNA Decay (NMD).

### Methods

#### Gene burden tests

To test for association between rare variants in genes and cancers of interest we performed simple burden tests where variants within genes are collapsed together. This is a powerful method if variants have similar effect sizes^20^. We consider the simplest type of burden test where genotypes are collapsed to a 0/1 variable based on whether samples carry a protein-truncating variant (PTV) in a specific gene. That is,*G*_*i*_ = 1 *if* 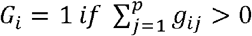 *and if* 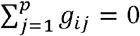 where *g*_*ij*_= 0, 1, 2 is the number of minor alleles observed for sample i at variant j, and p is the number of PTVs in the gene.

To apply the approach above, we fit logistic regression models in which the carrier status is the outcome variable, and the disease phenotype is a covariate. This method is described further elsewhere^21^.

When there is no family history information available and the cancer is prevalent in only one sex, e.g. ovarian cancer, the model used is:

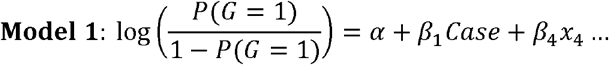

When there is no family history information available, but the cancer is prevalent in both sexes, e.g. pancreatic cancer, the model used is:

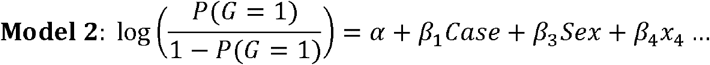

When family history information is available, e.g. for breast cancer, the model used is:

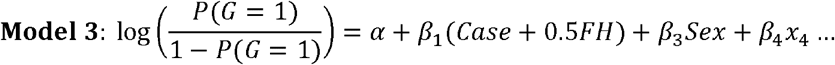

We tested for association using the Wald p-value associated with *β*_1_. Highly significant associations based on small counts may be unreliable, so we also conducted likelihood ratio tests for genes reaching exome-wide significance with case carriers ≤5.

For breast cancer, the PTV burden results were reported by Wilcox et al. (2023)^5^. For the other cancers with available family history information, i.e. bowel, lung, and prostate cancer, the results for the PTV burden association analysis are shown elsewhere^21^. Here we additionally present the PTV burden association results for the 7 other cancers without family history information.

#### Modelling effect sizes

A method to model the effect sizes associated with PTVs, and hence estimate the contribution of PTVs to the Familial Relative Risk of breast cancer, was described by Wilcox *et al* (2023)^5^. Here we generalise this method for any cancer in the UK Biobank, as well as extend the method to account for the joint distribution of multiple cancers. For breast cancer results here we use the UK Biobank data only, whereas the paper by Wilcox. et al (2023)^5^ included the BCAC data in the FRR calculations.

### Individual cancer model

For individual cancers, we assume a prior distribution for effect sizes (log-odds ratios) *f*(*β*| *α, η)* in which a proportion,*α*, of genes are associated with the cancer. For genes that are risk-associated, the prior distribution for the log-odds ratio is assumed to follow a negative exponential distribution. Thus

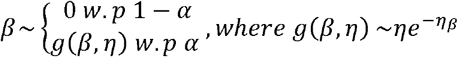

The effect size is assumed to be the same for all PTVs in a gene. Thus, the distribution is determined by the parameters *α* and *η*. These parameters can be fit, using maximum likelihood, from summary counts of the numbers of case and control PTV carriers in each gene and each sex. For cancers where family history is also available, the method cancer be extended to incorporate counts by family history. Details of the likelihood derivation are given in Supplementary Methods and Wilcox et al^5^. The estimates of *α* and *η* can be used the derive the posterior probability that each gene is risk-associated and the median predicted effect size. It can also be used to estimate the familial relative risk to first-degree relatives (*γ*)attributable to PTVs in all genes, under the assumption that the combined effect of PTVs in different genes is additive^5,22^. We also express this as an estimated proportion of the overall familial relative risk under the assumption that the overall FRR for each cancer is 2 (which is approximately true for all the cancers considered here^23,24^) and that the PTVs combine multiplicatively with other common genetic or familial factors.

To model the joint effect of two cancers, we extend the model to allow four categories of gene: genes associated with cancer 1 only, cancer 2 only, both cancers, or neither cancer, with proportions *α*_10_, *α*_01_, *α*_11_ and 1 − *α*_10_ − *α*_01_ − *α*_11_ respectively. The effect sizes for the two cancers, when associated, are again assumed to follow negatively exponential distributions. Thus:

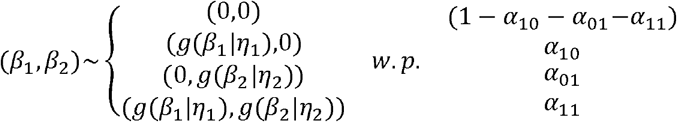

Where *g*(*β*_*1*_ |*η*_1_) ∼ *η*_1_ exp(− *η*_1_ *β*_*1*_), *g*(*β*_*2*_ |*η*_2_) ∼ *η*_2_ exp(− *η*_2_ *β*_*2*_)

There are thus five parameters to estimate: *α*_10_, *α*_01_, *α*_11_, *η*_1_ and *η*_2_.

For simplicity, we assume that the effect sizes (when both cancers are associated) *β*_*1*_ and *β*_*2*_ are uncorrelated. This is motivated by the fact that the data would be too limited to estimate this correlation in addition to the other parameters. Moreover, for the strongest known genes associated with multiple cancers (e.g. *ATM, BRCA1, BRCA2*), there is no clear correlation between the effect sizes for different cancers. To evaluate the evidence for overlap in the susceptibility genes for pairs of cancers, we performed likelihood ratio tests against the null hypothesis that the probabilities that genes are associated with each of the two cancers are independent, i.e. the odds ratio:

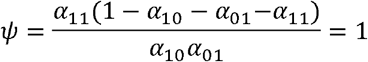

More details are provided in the Supplementary Methods.

## Results

### PTV burden results

The PTV burden results for 11 all cancers are summarised in Table 2. Association results for each cancer that were not reported previously^5,21^, for genes reaching P<0.001, can be found in Supplementary Tables 1 to 7. The corresponding Manhattan and QQ plots can be found in Supplementary Figures 1 to 14. A comparison of Wald and LRT P-values for genes reaching exome-wide significance from the Wald test and with case carriers ≤ 5 for each cancer are shown in Supplementary Table 8.

**Table 2.** Summary of results for PTV burden tests for 11 cancers. The table includes the number of genes reaching different significance thresholds as well as the list of genes that reach exome-wide significance and COSMIC TSGs with Wald test P<1×10-4 listed in ascending p-value order. For breast cancer, the results are from the meta-analysis of the UK Biobank and BCAC dataset as reported by Wilcox et al^5^.

| Cancer | Number of genes at a P-value threshold | | | Genes with $P < 2.5 \times 10^{-6}$ | COSMIC TSGs with $P < 1 \times 10^{-4}$ |
| --- | --- | --- | --- | --- | --- |
| | $P < 0.001$ | $P < 1 \times 10^{-4}$ | $P < 2.5 \times 10^{-6}$ | | |
| **Breast** | 30 | 9 | 6 | *BRCA2, BRCA1, CHEK2, PALB2, ATM, MAP3K1* | *BRCA2, BRCA1, CHEK2, PALB2, ATM, MAP3K1, LZTR1, BARD1* |
| **Prostate** | 35 | 8 | 3 | *BRCA2, CHEK2, ATM* | *BRCA2, CHEK2, ATM* |
| **Bowel** | 42 | 9 | 5 | *MSH6, MSH2, MLH1, APC, GAPDH* | *MSH6, MSH2, MLH1, APC, FLCN, SMAD4* |
| **Lung** | 46 | 8 | 0 | NA | *ARHGAP35, BIRC3* |
| **Pancreatic** | 100 | 38 | 8 | *ATM, MEN1, RCN2, YPEL3, SMC2, SEC14L3, GNG10, ZNF461* | *ATM, MEN1* |
| **Endometrial** | 80 | 26 | 5 | *MSH6, MLH1, ACRV1, STK32C, PSMC6* | *MSH6, MLH1, MSH2* |
| **Ovarian** | 115 | 35 | 9 | *BRCA2, BRCA1, IVD, JAML, KCNAB2, ZFP14, TMEM163, TMEM167A, NHEJ1* | *BRCA2, BRCA1, NFRSF14* |
| **Oesophagus** | 117 | 32 | 2 | *KNL1, IRF2BP2* | *KNL1, ATM, FUS* |
| **Kidney** | 89 | 27 | 6 | *PKD1, FGL2, TTC9, EXOC7, NCK2, TMEM174* | NA |
| **Bladder** | 72 | 27 | 8 | *DLX2, ZNF506, CDCP2, TMEM222, KDM1A, ARHGEF6, HR, NLRP10* | NA |
| **Malignant Melanoma** | 45 | 12 | 4 | *DCX, MED9, MRPL44, CDKN2A* | *CDKN2A* |

The cancer with the most exome-wide associations by the Wald test was ovarian cancer (9 genes), followed by pancreatic (8 genes) and bladder cancer (8 genes). We examined specifically tumour suppressor genes (TSGs) defined by COSMIC since previous analysis for breast cancer indicated that this category was highly enriched^5^. The cancer with the most COSMIC TSGs having P<1×10^−4^ was breast cancer (8 genes) followed by bowel cancer (6 genes). Among TSGs, *ATM* was associated at P<1×10^−4^ for breast, prostate, pancreatic and oesophagus cancer; *BRCA2* was associated at P<1×10^−4^ for breast, prostate, and ovarian cancer; and *MSH6, MSH2* and *MLH1* were associated at P<1×10^−4^ for bowel and endometrial cancer. No Cosmic TSGs had P<1×10^−4^ for kidney or bladder cancer. Cancers with >50 genes associated at P<0.001 include pancreatic, endometrial, ovarian, oesophagus, kidney, and bladder cancer. However, we note that for these cancers the number of cases was quite small and that for many of the associated genes, the number of case carriers was very low.

Of genes not listed in Table 2, of interest for ovarian cancer are associations at P<0.05 for 5 putative ovarian cancer genes: *MSH6* (p=0.00056), *BRIP1* (p=0.00055), *RAD51C* (p=0.028), *RAD51D* (p=0.00011) and *CHEK2* (p=0.00049).

### Empirical Bayes modelling

Table 3 and Figure 1 summarise the best-fitting models for each cancer. The cancer with the greatest estimated proportion of risk-associated genes was ovarian cancer (α=0.037, equivalent to 578 genes), followed by oesophageal cancer (α=0.030, 468 genes) and pancreatic cancer (α=0.019, 297 genes). In contrast, for kidney, bladder, and malignant melanoma α was estimated to be 0. The estimated exponential distributions for the log odds ratios are shown in Figure 1. The graph is least steep for bowel cancer, which has the greatest estimated median odds ratio. In contrast, the median odds ratio was lowest for prostate cancer, consistent with a lower proportion of higher-risk genes.

**Table 3.** Optimised values of *α* and *η* for each cancer, as well as the estimated heritability and contribution to familial relative risk. The %FRR estimates use posterior gene PTV frequency adjusted for CNV frequency. The genes with posterior>0.5 are listed in descending order.

| | $\alpha$ | $\eta$ | median OR | loglik | $\lambda$ | %FRR | Genes with posterior>0.9 | Genes with posterior>0.5 |
| --- | --- | --- | --- | --- | --- | --- | --- | --- |
| **Breast** | 0.0027 | 1.8 | 1.5 | 713.43 | 1.06 | 8.8 | *BRCA1, BRCA2, CHEK2, PALB2, ATM, MAP3K1* | *BRCA1, BRCA2, CHEK2, PALB2, ATM, MAP3K1* |
| **Prostate** | 0.020 | 6.2 | 1.1 | 37.73 | 1.01 | 1.08 | *BRCA2, CHEK2, ATM* | *BRCA2, CHEK2, ATM* |
| **Bowel** | 0.0016 | 1.2 | 1.8 | 140.67 | 1.05 | 6.7 | *MSH2, MSH6, MLH1, APC* | *MSH2, MSH6, MLH1, APC, GAPDH* |
| **Lung** | 0.0035 | 7.4 | 1.1 | 0.04 | 1.00 | 0.1 | *NA* | *NA* |
| **Pancreas** | 0.019 | 2.8 | 1.3 | 7.26 | 1.08 | 10.6 | *ATM* | *ATM* |
| **Endom** | 0.0014 | 1.3 | 1.7 | 88.05 | 1.12 | 16.5 | *MSH6* | *MSH6* |
| **Ovarian** | 0.037 | 2.4 | 1.3 | 89.26 | 1.37 | 45.9 | *BRCA2, BRCA1* | *BRCA2, BRCA1, IVD, SLC35E4, REXO5* |
| **Oesophagus** | 0.030 | 3.6 | 1.2 | 0.92 | 1.04 | 6.2 | *NA* | *NLRP12, ATM* |
| **Kidney** | 0.00 | 4.4 | 1.2 | 0.00 | 1.00 | 0.0 | *NA* | *NA* |
| **Bladder** | 0.00 | 2.5 | 1.3 | 0.00 | 1.00 | 0.0 | *NA* | *NA* |
| **Malignant Melanoma** | 0.00 | 5.6 | 1.1 | 0.00 | 1.00 | 0.0 | *NA* | *NA* |

**Table 4.**
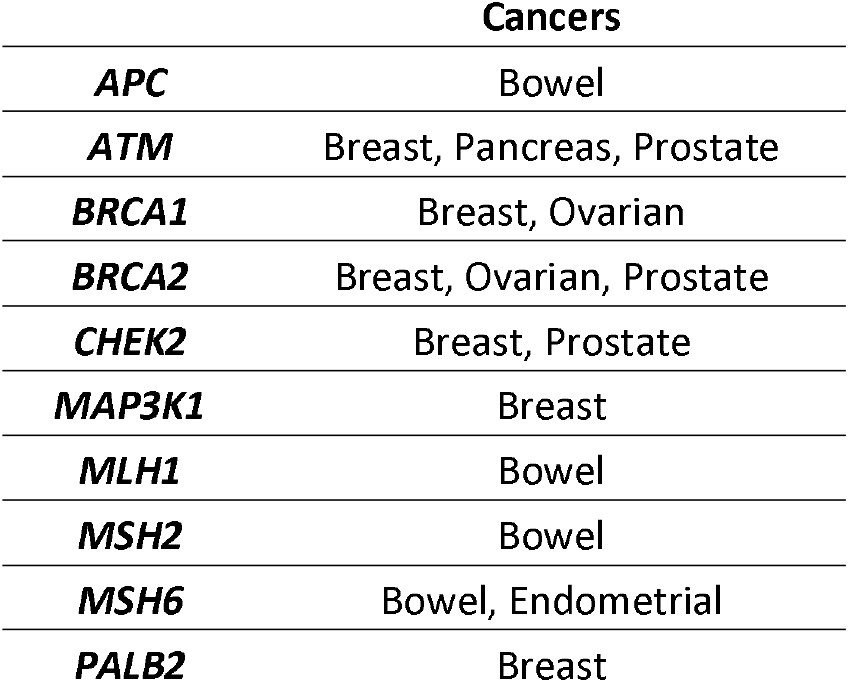
Genes with posterior>0.8 for at least 1 cancer. The cancer columns are the cancers which had a posterior probability >0.8 of the gene being associated with the cancer.

**Figure 1.**
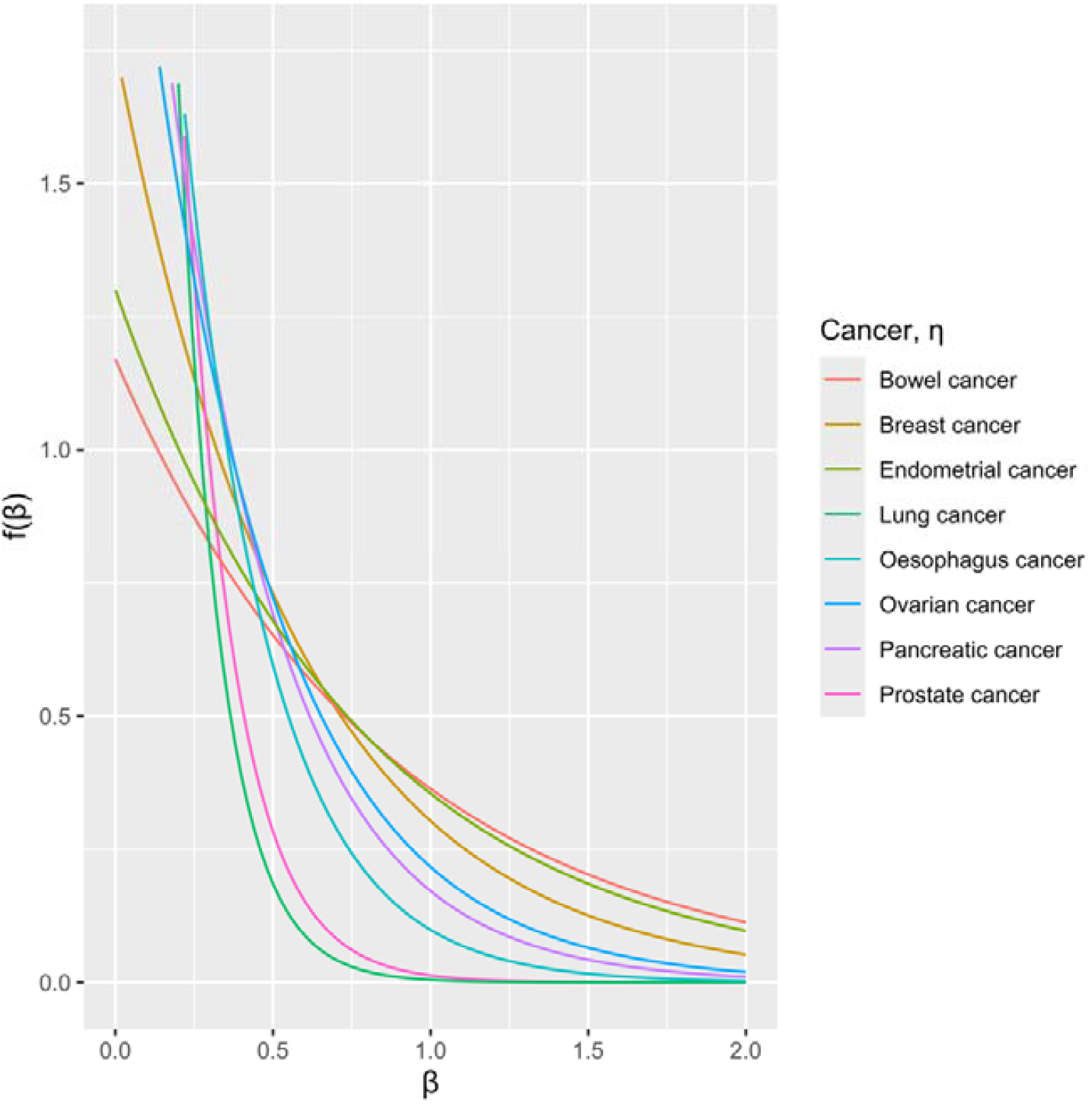
The exponential distributions for the log-odds ratio of associated genes for each cancer. This uses the optimised value of *η*, where *gβ, η*∼ *ηe - η β*

In terms of overall contribution to the FRR, the highest proportion was for ovarian cancer (45.9%) followed by endometrial cancer (16.5%; Table 3).

Based on the best-fitting models, we computed the posterior probability for each gene to be associated with each cancer (Supplementary Tables 9 to 16). For breast cancer, there were 6 genes with posterior probability >0.9: *BRCA2, BRCA1, PALB2, CHEK2, ATM* and *MAP3K1* (Supplementary Table 9). *BRCA2* and *BRCA1* also reached this level for ovarian cancer, *BRCA2, ATM* and *CHEK2* for prostate cancer, and *ATM* for pancreatic cancer. For bowel cancer, there were 4 genes with posterior probability>0.9: the mismatch repair genes *MLH1, MSH6* and *MSH2*, as well as *APC* (Supplementary Table 10). *MSH6* also reached this level for endometrial cancer. For lung, oesophagus, kidney, and bladder cancer, as well as malignant melanoma, there were no genes with posterior probability >0.9.

### Joint cancer models

We next considered models incorporating pairs of cancers (Figures 2 and 3). The strongest evidence for overlap was found for breast-prostate (likelihood ratio P=1.5×10^−9^), breast-ovarian (P=2.1×10^−8^), bowel-endometrial (P=3.0×10^−8^) and breast-pancreatic (P=2.3×10^−5^). Associations with p<0.001 were additionally observed for prostate-ovarian and prostate-pancreas, with weaker evidence of overlap for breast-bowel, breast-lung, breast-oesophagus and lung-pancreas. More detailed results for pairs with P<0.01 are shown in Table 5 and for all cancer pairs in Supplementary Table 17.

**Table 5.** Estimated values of *α*_10_, *α*_11_, *α*_11_, *η*_1_ and *η*_2_, as well as LRT p-value from comparing enrichment to cancer 2 given cancer 1 to the cancers being independent. Method refers to the joint cancer model method used as described in the method section. Results here have LRT p-value<0.01 and are sorted by ascending LRT p-value. *α*_10_ = *P(C*1 *n C*2^′^),*α*_01_ = *P(C*1^′^ *n C*2) *and α*_11_ = *P(C*1^′^ *n C*2).

| Cancer 1 | Cancer 2 | Method | $\alpha_{10}$ | $\alpha_{01}$ | $\alpha_{11}$ | $\eta_1$ | $\eta_2$ | LRT |
| --- | --- | --- | --- | --- | --- | --- | --- | --- |
| Breast | Prostate | 1 | 0.00000 | 0.00000 | 0.00220 | 1.49 | 2.51 | 1.5E-09 |
| Breast | Ovarian | 1 | 0.00000 | 0.00000 | 0.00270 | 1.77 | 1.23 | 2.1E-08 |
| Bowel | Endom | 2 | 0.00020 | 0.00000 | 0.00140 | 1.13 | 0.93 | 3.0E-08 |
| Breast | Pancreas | 1 | 0.00000 | 0.00000 | 0.00230 | 1.62 | 1.57 | 2.3E-05 |
| Prostate | Ovarian | 1 | 0.00000 | 0.00340 | 0.01700 | 6.04 | 2.06 | 0.00015 |
| Prostate | Pancreas | 1 | 0.00000 | 0.00000 | 0.00840 | 4.56 | 2.17 | 0.00028 |
| Breast | Lung | 1 | 0.00013 | 0.00026 | 0.00150 | 1.52 | 6.00 | 0.0014 |
| Breast | Bowel | 1 | 0.00053 | 0.00027 | 0.00160 | 1.51 | 1.00 | 0.0035 |
| Pancreas | Ovarian | 1 | 0.00000 | 0.00000 | 0.03000 | 3.09 | 2.25 | 0.0054 |
| Breast | Oesophagus | 1 | 0.00000 | 0.00980 | 0.00240 | 1.64 | 2.95 | 0.0062 |
| Lung | Pancreas | 1 | 0.00000 | 0.00000 | 0.00460 | 5.93 | 2.05 | 0.0069 |

**Figure 2.**
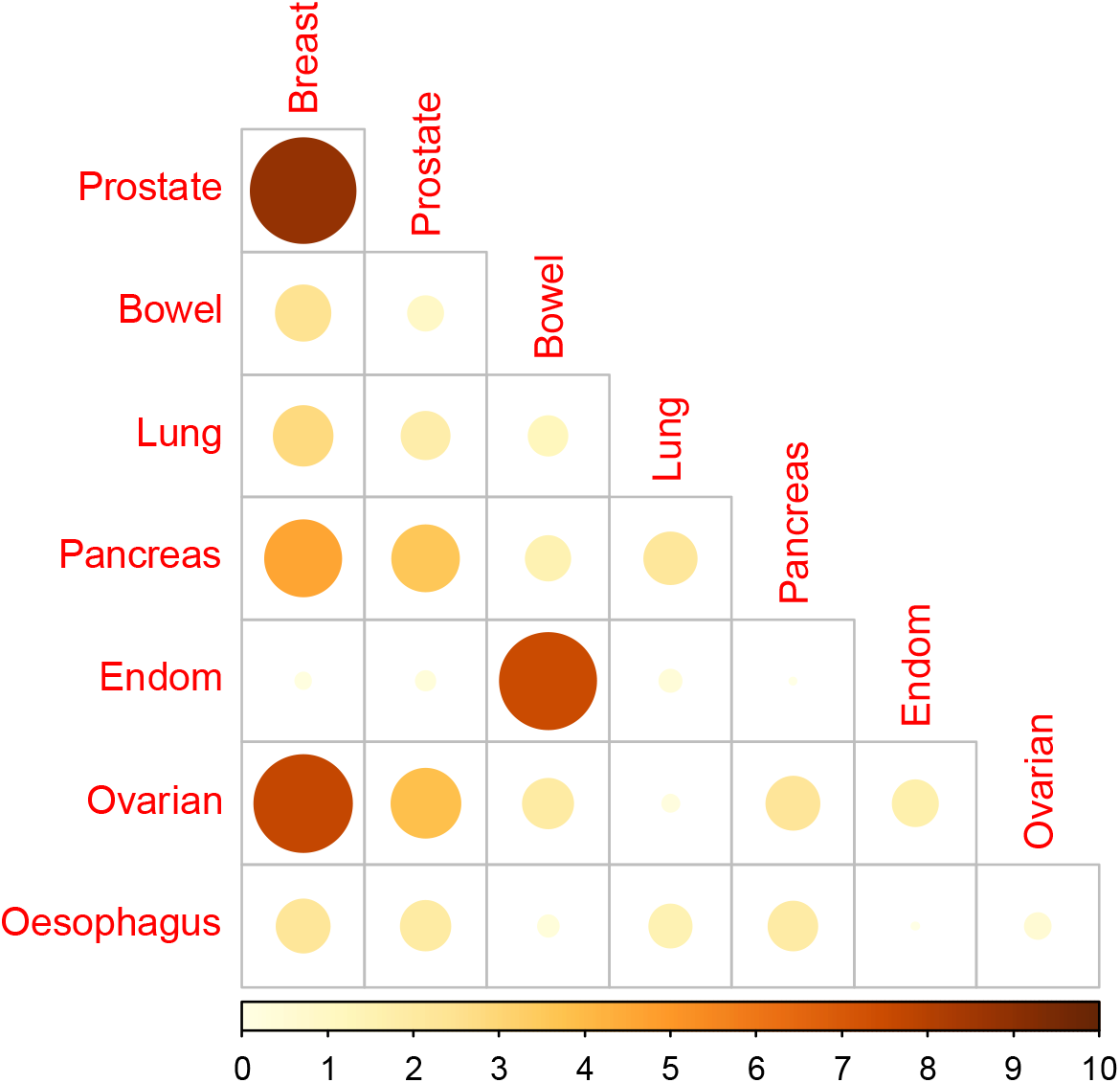
-log_10_(P-value) from the LRT of testing the model of enrichment to cancer 2 given cancer 1 to when the cancers are independent.

**Figure 3.**
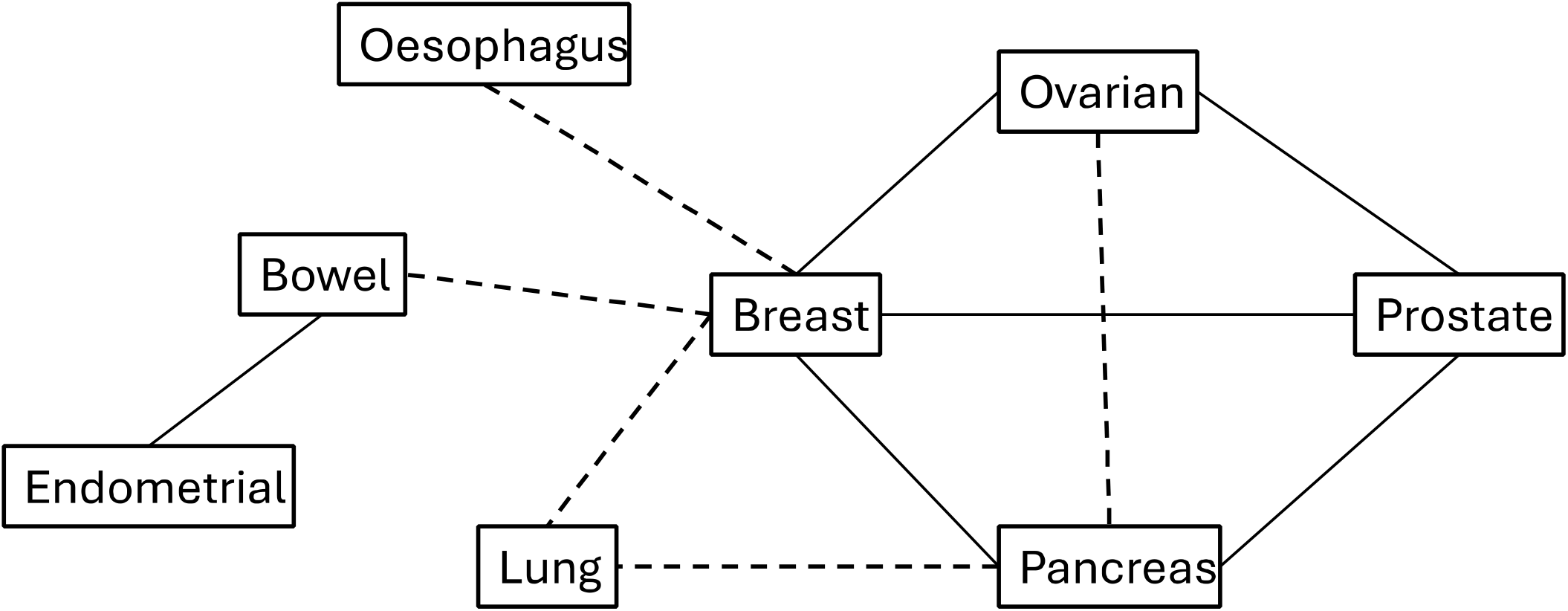
| Diagram showing cancer pairs with LRT p≤0.001 in bold and LRT 0.01 ≤ p< 0.001 in dashed.

We note that of the cancer pairs with P<0.001, the breast-prostate, breast-ovarian, breast-pancreatic, prostate-pancreatic, and pancreatic-ovarian pairs all estimated *α*_10_ = *α*_01_ = 0, i.e. complete overlap of associated genes. Genes with posterior>0.8 of being associated with both cancers for at least 1 cancer pair were *APC, ATM, BAP1, BRCA1, BRCA2, CHEK2, MAP3K1, MLH1, MSH2, MSH6* and *PALB2* (Supplementary Table 18). *ATM* was associated with the most cancer pairs with posterior >0.8 (9 pairs). Six of these genes are established genes for one or more of the cancers, the exception being BAP1 which had a posterior probability 0.824 in the breast-prostate model. In the breast-ovarian model, the gene with the next highest posterior probability (after those identified above) was *NHEJ1* (0.300), followed by the established risk genes *RAD51D* (0.282) and *BRIP1* (0.259) (Supplementary Table 19). *NHEJ1* PTVs were associated with an odds ratio of 2.70 (1.30, 5.61), p=0.0079 for breast cancer and 17.67 (5.36, 58.24), p=2.37×10^−6^for ovarian cancer.

For bowel and endometrial cancer, the best joint model estimated *α*_1_ = 0.00020 (≈3 genes), *α*_2_ = 0 and *α*_3_ =0.00140 (≈22 genes), i.e. consistent with subsets of genes associated with both cancers and with bowel cancer only, but none associated with endometrial cancer alone. There were 3 genes with a posterior probability>0.9 of being associated with both cancers: *MSH6* (1.00), *MLH1* (0.98) and *MSH2* (0.98) (Supplementary Table 21). COSMIC TSGs with posterior>0.5 additionally include *APC* (0.52). The genes with the highest posterior probability of being associated with bowel cancer alone were *APC* (0.48) and *GAPDH* (0.48).

## Discussion

We used a large exome-sequencing dataset to describe the pattern of gene-based associations across 11 cancer types. For the cancers with results not previously reported elsewhere, we found exome-wide significant genes for pancreatic, endometrial, ovarian, oesophagus, kidney, and bladder cancer as well as for malignant melanoma.

Our results additionally highlight the potential value of combining information for multiple cancers. We found strong evidence for overlap in susceptibility genes between several cancer pairs. This information can strengthen the evidence for susceptibility to one cancer by borrowing information from the other cancer, in effect improving the prior. Thus, we found the data to be consistent with an essentially complete overlap of susceptibility genes for breast, ovarian, pancreatic and prostate cancer. While this is not surprising given that several of the known susceptibility genes are associated with several of these cancers (though not to the same extent), the results highlight some additional putative genes.

Of particular interest was *NHEJ1*, which reached exome-wide significance in ovarian cancer analysis and showed evidence of association with breast cancer. The latter association would have been missed if considered in isolation. *NHEJ1* encodes a DNA repair factor essential for non-homologous end joining (NHEJ). The association for PTVs in this gene is novel but consistent with NHEJ being defective in a large proportion of ovarian cancers^25^. Ovarian cancer cells defective in NHEJ have also been shown to be resistant to PARP inhibition^25^, which might indicate some relevance for therapeutic approaches. We also found stronger evidence for *BAP1* in the joint analysis of breast and prostate cancer. *BAP1* PTVs have been associated with a range of cancers including cutaneous and uveal melanoma, kidney cancer, mesothelioma and basal cell carcinoma. Kidney (P=0.059) and bladder cancer (P=0.047), as well as ovarian (P=0.033) and pancreatic cancer (P=0.022), also showed some evidence of association in this dataset (Supplementary Table 30, Supplementary Figure 15).

Beyond the breast, ovarian, pancreatic and prostate associations, and the expected overlap between bowel and endometrial cancer, driven by the mismatch repair genes, there were also significant though weaker overlaps between breast cancer and several other cancers. This is partly driven by *ATM*, which is known to be associated with multiple cancers and had a posterior probability >0.8 of being associated with both cancers in 9 pairs (involving bowel, lung and oesophageal cancer in addition to breast, prostate, ovary and pancreas). The best fitting models for lung cancer in combination with breast, ovarian, pancreatic and prostate cancer are all consistent with most susceptibility genes for these cancers also being associated with lung cancer, albeit to a smaller extent.

The results confirm the striking enrichment of tumour suppressor and DNA repair genes among cancer susceptibility genes. Thus, of the 19 genes with a posterior probability >0.5 in any analyses, 12 are known tumour suppressor genes and/or involved in DNA repair (including all 11 genes with a posterior probability >0.8). In terms of the contribution of PTVs to the FRR of each cancer, the proportion was estimated to be greatest for ovarian cancer (46%), followed by endometrial cancer (16.5%). The contribution was estimated to be 0% for kidney and bladder cancer and malignant melanoma. This is clearly not exactly true: *CDKN2A* and *CDK4* PTVs are associated with malignant melanoma^26,27^, and VHL is associated with kidney cancer^28^. (*CDKN2A* did reach exome-wide significance for association with melanoma in this dataset, using a Wald test, but this is likely to have been exaggerated, the likelihood ratio p-value was 0.00041). *CDK4* had 0 melanoma case carriers, while *VHL* had 1 kidney case carrier and the OR was non-significant (3.5 (0.49, 25.4), P=0.21). Larger datasets should allow more precise estimates for the associations between these genes and cancer risk to be derived. Notwithstanding these uncertainties, the estimated contribution to the FRR, for all cancers, was largely attributable to the known genes. This strongly suggests that the contribution of additional genes is likely to be small and that most “missing” heritability is likely to reflect non-coding variation.

Similar to the breast cancer results reported previously, there was an excess of associations at P<0.001 across the cancers, indicating that further genes should be identifiable in larger datasets. We also note that for many cancers, the sample size is small, resulting in large standard errors and wide confidence intervals for many associated genes. Estimated risk estimates for novel associated genes may also be over-estimated due to the ‘winners curse’^29^. Further replication in larger datasets will therefore be necessary to confirm associations and to provide more precise risk estimates for variants in associated genes.

Our analyses have some limitations. We restricted our analyses to PTVs. For the known genes, most of the effect is driven by PTVs, and the assumption that PTVs confer similar risks is a plausible simplification. In principle, the analyses could be extended to missense, or other, coding variants, but this would require the model to be extended to incorporate variation in effect size. Our multicancer analyses have thus far been restricted to pairs of cancers but could logically be extended to larger sets of cancers. This would provide a more rational model, defining a single set of parameters and posterior probabilities. However, it would require a larger number of parameters to be estimated simultaneously. The model also assumed a particular prior distribution of effect sizes, in which a proportion of genes are associated with risk -an example of a spike-and-slab prior^30^. This is somewhat analogous to the approach used in some GWAS analyses, for example, LDpred2^31^. The model also only accounts for rare variants associated with an increased risk and does not account for protective alleles. We note, however, that all the 47 exome-wide significant associations were positive, so this appears a reasonable simplification given the available data. The model is simplistic and may not reflect the true underlying distribution for all cancers, however, the model provides a systematic approach to identify and rank genes worthy of exploration in larger targeted sequencing and functional experiments.

In conclusion, we have developed an approach to estimating the genome-wide contribution of the burden of rare coding variants to the heritability of cancer, considering 11 cancers in the UK Biobank. We have shown significant clustering of cancer types, including breast, ovarian, prostate and pancreatic cancer, with a large enrichment of tumour suppressor and DNA repair genes among cancer susceptibility genes. The estimated contribution to the FRR, for all cancers, was largely attributable to the known genes. This strongly suggests that the contribution of additional genes is likely to be small and that most “missing” heritability is likely to reflect non-coding variation.

## Data Availability

Requests for access to UK Biobank data should be made to the UK Biobank Access Management Team.

## Acknowledgements

Quality control of the UK Biobank sequencing data has been funded by the Medical Research Council (unit programs: MC_UU_12015/2, MC_UU_00006/2). The research has been conducted using the UK Biobank Resource under Application Number 28126. N.W. was supported by the International Alliance for Cancer Early Detection, an alliance between Cancer Research UK (C14478/A29329), the Canary Center at Stanford University, the University of Cambridge, OHSU Knight Cancer Institute, University College London, and the University of Manchester. J.D. was supported by core funding from the NIHR Cambridge Biomedical Research Centre (NIHR203312). X.W. and J.P.T. were supported by Cancer Research UK (PPRPGM-Nov20\100002 and PRCPJT-May21\100006).

## Author Contributions

D.F.E. supervised this work and directed the overall analysis. N.W. performed the statistical analysis. N.W., E.J.G., J.P.T., and J.D.P. developed the bioinformatics and computational pipelines. X.Y. and J.D. acquired data and X.Y. extracted cancer phenotypes. N.W. and D.F.E. drafted the manuscript. All authors reviewed and approved the paper.

## Data and Code Availability

Requests for access to UK Biobank data should be made to the UK Biobank Access Management Team. Quality Control filtering of vcf files was performed using vcftools v0.1.15, bcftools v1.9, picard v2.22.2 and plink v1.90b, as outlined in the methods. Variants were annotated using Ensembl Variant Effect Predictor v101 with assembly GRCh38. The code for each software is available at the website of each package. Data manipulation and analysis were performed using R-4.3.3 with packages clusterProfiler (4.2.2), data.table (1.14.2), dplyr (1.0.9), dbplyr (2.5.0), gtools (3.9.5), HGNChelper (0.8.9), SKAT (2.2.5), tibble (3.2.1) and tidyr (1.3.1). Plots were created using additional packages ggplot2 (3.5.1) and ggrepel (0.9.5). The code for each of the R packages can be found in their associated vignettes.

