## Supplementary Material for "The contribution of coding variants to the heritability of multiple cancer types using UK Biobank whole-exome sequencing data"

### Supplementary Methods

##### Individual cancer model

To derive the likelihood function we stratify the number of carriers by sex, family history, and case/control status.

|  | **FEMALE** | | | | | **MALE** | | | | |
| --- | --- | --- | --- | --- | --- | --- | --- | --- | --- | --- |
|  | Control | | Case | |  | Control | | Case | |  |
|  | FH 0 | FH 1 | FH 0 | FH 1 |  | FH 0 | FH 1 | FH 0 | FH 1 |  |
| **Non-carrier** | $N_{F0}-n_{F0j}$ | $N_{F1}-n_{F1j}$ | $N_{F2}-n_{F2j}$ | $N_{F3}-n_{F3j}$ | $N_{F}-n_{F}$ | $N_{M0}-n_{M0j}$ | $N_{M1}-n_{M1j}$ | $N_{M2}-n_{M2j}$ | $N_{M3}-n_{M3j}$ | $N_{M}-n_{M}$ |
| **Carrier** | $n_{F0j}$ | $n_{F1j}$ | $n_{F2j}$ | $n_{F3j}$ | $n_{F0j}+n_{F1j}{+n}_{F2j}+n_{F3j}=n_{F}$ | $n_{M0j}$ | $n_{M1j}$ | $n_{M2j}$ | $n_{M3j}$ | $n_{M0j}+n_{M1j}{+n}_{M2j}+n_{M3j}=n_{M}$ |
|  | $N_{F0}$ | $N_{F1}$ | $N_{F2}$ | $N_{F3}$ | $N_{F0}+N_{F1}+N_{F2}+N_{F3}=N_{F}$ | $N_{M0}$ | $N_{M1}$ | $N_{M2}$ | $N_{M3}$ | $N_{M0}+N_{M1}+N_{M2}+N_{M3}=N_{M}$ |

For each sex, the number of carriers in each stratum can be modelled by a multinomial distribution with a probability mass function:

$P\left( n_{oj},n_{1j}, n_{2j}, n_{3j}|n_{j},\beta_{j} \right)=\frac{\prod_{k=0}^{3} \left( N_{k}\frac{1}{2}\left( ke^{\beta_{j}}+2-k \right) \right)^{n_{kj}}}{\left( \sum_{k=0}^{3} N_{k}\frac{1}{2}\left( ke^{\beta_{j}}+2-k \right) \right)^{n_{j}}}$

Defining $\gamma_{Fk}=log(\frac{N_{Fk}}{N_{F0}})$, $\gamma_{Mk}=log(\frac{N_{Mk}}{N_{M0}})$, and multiplying the probabilities for males and females, this simplifies to:

$$P\left( n_{oj},n_{1j}, n_{2j}, n_{3j}|n_{j},\beta_{j} \right)=C\frac{\prod_{k=0}^{3} \left( \frac{1}{2}\left( ke^{\beta_{j}}+2-k \right) \right)^{n_{Fkj}}\prod_{k=0}^{3} \left( \frac{1}{2}\left( ke^{\beta_{j}}+2-k \right) \right)^{n_{Mkj}}}{\left( \sum_{k=0}^{3} e^{\gamma_{F_{k}}}\frac{1}{2}({ke}^{\beta_{j}}+2-k) \right)^{n_{Fj}}\left( \sum_{k=0}^{3} e^{\gamma_{M_{k}}}\frac{1}{2}({ke}^{\beta_{j}}+2-k) \right)^{n_{Mj}}}$$

Where $C=\left( \begin{matrix} n_{Fj} \\ \begin{matrix} \begin{matrix} n_{F0j} & n_{F1j} \end{matrix} & \begin{matrix} n_{F2j} & n_{F3j} \end{matrix} \end{matrix} \end{matrix} \right)\left( \begin{matrix} n_{Mj} \\ \begin{matrix} \begin{matrix} n_{M0j} & n_{M1j} \end{matrix} & \begin{matrix} n_{M2j} & n_{M3j} \end{matrix} \end{matrix} \end{matrix} \right)e^{\sum_{k=0}^{3} \gamma_{Fk}n_{Fkj}+\sum_{k=0}^{3} \gamma_{Mk}n_{Mkj}}$ is independent of the prior distribution.

The likelihood is integrated over the prior distribution to give the likelihood to be maximised:

$$L(\alpha,\eta)\propto\prod_{j=1}^{J} \int\frac{\prod_{k=0}^{3} \left( \frac{1}{2}\left( ke^{\beta_{j}}+2-k \right) \right)^{n_{Fkj}}\prod_{k=0}^{3} \left( \frac{1}{2}\left( ke^{\beta_{j}}+2-k \right) \right)^{n_{Mkj}}}{\left( \sum_{k=0}^{3} e^{\gamma_{F_{k}}}\frac{1}{2}({ke}^{\beta_{j}}+2-k) \right)^{n_{Fj}}\left( \sum_{k=0}^{3} e^{\gamma_{M_{k}}}\frac{1}{2}({ke}^{\beta_{j}}+2-k) \right)^{n_{Mj}}}f\left( \beta_{j}|\alpha,\eta\right)d\beta_{j}$$

Where $f(\beta_{j}|\alpha,\eta)$ is the prior distribution on $\beta_{j}$.

Writing$L_{j}\left( \beta_{j} \right)=\left( \sum_{k=0}^{3} e^{\gamma_{F_{k}}} \right)^{n_{Fj}}\left( \sum_{k=0}^{3} e^{\gamma_{M_{k}}} \right)^{n_{Mj}}\frac{\prod_{k=0}^{3} \left( \frac{1}{2}\left( {ke}^{\beta_{j}}+2-k \right) \right)^{n_{Fkj}}\prod_{k=0}^{3} \left( \frac{1}{2}{(ke}^{\beta_{j}}+2-k) \right)^{n_{Mkj}}}{\left( \sum_{k=0}^{3} e^{\gamma_{F_{k}}}\frac{1}{2}\left( \left( {ke}^{\beta_{j}}+2-k \right) \right) \right)^{n_{Fj}}\left( \sum_{k=0}^{3} e^{\gamma_{M_{k}}}\frac{1}{2}{(ke}^{\beta_{j}}+2-k) \right)^{n_{Mj}}}$

$L\left( \alpha,\eta\right)\propto\prod_{j=1}^{J} \int L_{j}\left( \beta_{j} \right)f\left( \beta_{j}|\alpha,\eta\right)d\beta_{j}=\prod_{j=1}^{J} \left( 1-\alpha+\alpha\int L_{j}\left( \beta_{j} \right)g\left( \beta_{j}|\eta\right)d\beta_{j} \right)=\prod_{j=1}^{J} \left( 1-\alpha+\alpha L_{*j} \right)$.

The posterior probability a gene is associated, given optimised estimates of $\alpha$and $\eta$ is: $P\left( \beta_{j} | Data \right)=\frac{\alpha\int L_{j}(\beta_{j})g\left( \beta_{j}|\eta\right)d\beta_{j}}{1-\alpha+\alpha\int L_{j}(\beta_{j})g\left( \beta_{j}|\eta\right)d\beta_{j}}=\frac{\alpha L_{*j}}{1-\alpha+\alpha L_{*j}}$.

The posterior mean $\beta_{j}$ is: $\frac{\int{\beta_{j}L}_{j}(\beta_{j})g\left( \beta_{j}|\eta\right)d\beta_{j}}{L_{*j}}$.

And the posterior mean relative risk $e^{\beta_{j}}$ is: $\frac{\int{e^{\beta_{j}}L}_{j}(\beta_{j})g\left( \beta_{j}|\eta\right)d\beta_{j}}{L_{*j}}$.

For gene j with aggregate PTV frequency, $p_{j}$, associated with relative risk $e^{\beta_{j}}$, the FRR is $\lambda_{j}=1+\frac{p_{j}{(e^{\beta_{j}}-1)}^{2}}{{(2p_{j}\left( e^{\beta_{j}}-1 \right)+1)}^{2}}$

Using control and case data, we estimate the allele frequency based on the posterior distribution of the relative risk: $p_{Bj}(\beta_{j})=\frac{n_{F0j}+n_{F1j}+n_{F2j}+n_{F3j}}{2(N_{F0}+N_{F1}+{e^{\beta_{j}}(N}_{F2}+N_{F3})}$.

Hence $\lambda_{jB}=1+\frac{\alpha}{1-\alpha+\alpha L_{*j}}\int L_{j}(\beta_{j})g\left( \beta_{j}|\eta\right)\frac{p_{jB}(\beta_{j}){(e^{p_{jB}(\beta_{j})}-1)}^{2}}{{(2p_{jB}(\beta_{j})\left( e^{p_{jB}(\beta_{j})}-1 \right)+1)}^{2}}d\beta_{j}$

The total FRR over all genes, assuming an additive model, is given by: $\hat{\lambda}_{TOT} =1+\sum_{j=1}^{J} (\lambda_{j}-1)$.

Assuming that the PTVs combine multiplicatively with other genetic or familial factors, and an overall FRR of 2, the percentage contribution of each gene to the overall FRR is: $100\times\frac{log\left( \hat{\lambda}_{j} \right)}{log(2)}$ and the total contribution of PTVs in all genes is: $100\times\frac{log\left( \hat{\lambda}_{TOT} \right)}{log(2)}$.

These equations simplify if there is no recorded family history information. In this case, the multinomial distribution simplifies to a binomial distribution. We set $N_{F1}=N_{F3}=N_{M1}=N_{M3}=0$. Therefore for each sex: $P\left( n_{oj}, n_{2j}|n_{j},\beta_{j} \right)\propto\frac{\left( N_{0} \right)^{n_{0j}}\left( N_{2}e^{\beta_{j}} \right)^{n_{2j}}}{\left( N_{0}+N_{2}e^{\beta_{j}} \right)^{n_{j}}}$

Defining $\gamma_{F2}=log(\frac{N_{F2}}{N_{F0}})$, $\gamma_{M2}=log(\frac{N_{M2}}{N_{M0}})$, as above, and multiplying the probabilities for males and females, this simplifies to:

$P\left( n_{oj}, n_{2j}|n_{j},\beta_{j} \right)=C\frac{\left( e^{\gamma_{F2}}e^{\beta_{j}} \right)^{n_{F2j}}\left( e^{\gamma_{M2}}e^{\beta_{j}} \right)^{n_{M2j}}}{\left( 1+e^{\gamma_{F2}}e^{\beta_{j}} \right)^{n_{Fj}}\left( 1+e^{\gamma_{M2}}e^{\beta_{j}} \right)^{n_{Mj}}}$. Here C=$\left( \begin{matrix} n_{Fj} \\ n_{F2j} \end{matrix} \right)\left( \begin{matrix} n_{Mj} \\ n_{M2j} \end{matrix} \right)$.

Therefore, $L(\alpha,\eta)\propto\prod_{j=1}^{J} \int\frac{\left( N_{F0} \right)^{n_{F0j}}\left( N_{F2}e^{\beta_{j}} \right)^{n_{F2j}}\left( N_{M0} \right)^{n_{M0j}}\left( N_{M2}e^{\beta_{j}} \right)^{n_{M2j}}}{\left( 1+e^{\gamma_{F2}}e^{\beta_{j}} \right)^{n_{Fj}}\left( 1+e^{\gamma_{M2}}e^{\beta_{j}} \right)^{n_{Mj}}}f\left( \beta_{j}|\alpha,\eta\right)d\beta_{j}$

##### Joint cancer model, method 1

We now consider two cancers and assume distributions for effect sizes in which a proportion, $\alpha_{1}$, of genes are associated with cancer 1, and $\alpha_{2}$ are associated with cancer 2. The log-relative risk for cancer 1, $\beta_{1}$, has a density of the form $f(\beta_{1}|\alpha_{1},\eta_{1})$ and the log-relative risk for cancer 2, $\beta_{2}$, has a density of the form $f(\beta_{2}|\alpha_{2},\eta_{2})$. We assume that the prior probabilities can be correlated (i.e., the probability that a gene is a risk for cancer 2 is dependent on whether it is a gene for cancer 1).

There are now 4 combinations for a gene j:

|  | | $\boldsymbol{Cancer 1,}\boldsymbol{\beta}_{\boldsymbol{1}\boldsymbol{j}}$ | |
| --- | --- | --- | --- |
|  |  | **0 (not associated)**  $w.p$ $\boldsymbol{(1-}\boldsymbol{\alpha}_{\boldsymbol{1}}\boldsymbol{)}$ | $\boldsymbol{g(}\boldsymbol{\beta}_{\boldsymbol{1}}\boldsymbol{\vert\eta)}$ **(associated)**  $w.p$ $\boldsymbol{\alpha}_{\boldsymbol{1}}$ |
| $\boldsymbol{Cancer 2,}\boldsymbol{\beta}_{\boldsymbol{2}\boldsymbol{j}}$ | **0 (not associated)**  $w.p$ $\boldsymbol{(1-}\boldsymbol{\alpha}_{\boldsymbol{2}}\boldsymbol{)}$ | (0,0)  $w.p$ $(1-\alpha_{10}-\alpha_{01}{-\alpha}_{11})$ | ($g(\beta_{1}\vert\eta_{1}),0)$  $w.p$ $\alpha_{10}$ |
|  | $\boldsymbol{g(}\boldsymbol{\beta}_{\boldsymbol{2}}\boldsymbol{\vert}\boldsymbol{\eta}_{\boldsymbol{2}}\boldsymbol{)}$ **(associated)**  $w.p \boldsymbol{\alpha}_{\boldsymbol{2}}$ | (0, $g(\beta_{2}\vert\eta_{2}))$  $w.p \alpha_{01}$ | ($g(\beta_{1}\vert\eta_{1}),g(\beta_{2}\vert\eta_{2})$)  $w.p \alpha_{11}$ |

i.e.

$(\beta_{1},\beta_{2})\sim\left\{ \begin{matrix} (0,0) w.p. (1-\alpha_{10}-\alpha_{01}{-\alpha}_{11}) \\ (g(\beta_{1}|\eta_{1}),0) w.p. \alpha_{10} \\ (0, g(\beta_{2}|\eta_{2})) w.p \alpha_{01} \\ (g(\beta_{1}|\eta_{1}),g(\beta_{2}|\eta_{2})) w.p. \alpha_{11} \end{matrix} \right.$

Where $g\left( \beta_{1} | \eta_{1} \right)\sim\eta_{1}\exp\left( -\eta_{1}\beta_{1} \right), g\left( \beta_{2} | \eta_{2} \right)\sim\eta_{2}\exp\left( -\eta_{2}\beta_{2} \right),$

For simplicity, we assume that the effect sizes $\beta_{1}$and $\beta_{2}$ are uncorrelated.

There are five parameters to estimate: $\alpha_{10}$, $\alpha_{01}$, $\alpha_{11}$, $\eta_{1}$, and $\eta_{2}$.

We can calculate $\alpha_{1}=\alpha_{10}+\alpha_{11}$ and $\alpha_{2}=\alpha_{01}+\alpha_{11}$. We note $\alpha_{1}, \alpha_{2}, \eta_{1}$, and $\eta_{2}$calculated here may differ to the values from optimising the individual cancer models.

The odds ratio $\alpha_{11}(1-\alpha_{10}-\alpha_{01}-\alpha_{11})/(\alpha_{10}\alpha_{01})$ represents the degree of enrichment of susceptibility to cancer 2 given cancer 1 (or vice versa).

The likelihood can be written as:

$$L\left( \alpha_{10},\alpha_{01},\alpha_{11},\eta_{1},\eta_{2} \right)\propto\prod_{j=1}^{J} \iint L_{j}\left( \beta_{1j},\beta_{2j} \right)f\left( \beta_{1j},\beta_{2j} | \alpha_{10},\alpha_{01},\alpha_{11} ,\eta_{1},\eta_{2} \right)d\beta_{1j}d\beta_{2j}=\prod_{j=1}^{J} \int L_{j}\left( \beta_{1j} \right)f\left( \beta_{1j} | \alpha_{1}\eta\right)d\beta_{1j}\int L_{j}\left( \beta_{2j} \right)f\left( \beta_{2j} | \alpha_{2}\eta\right)d\beta_{2j}=\prod_{j=1}^{J} \left( \left( 1-\alpha_{10}-\alpha_{01}-\alpha_{11} \right)+\alpha_{10}\int L_{j1}\left( \beta_{1j} \right)g\left( \beta_{1j}|\eta_{1} \right)d\beta_{1j}+\alpha_{01}\int L_{j2}\left( \beta_{2j} \right)g\left( \beta_{2j}|\eta_{2} \right)d\beta_{1j}+\alpha_{11}\int L_{j}\left( \beta_{1j} \right)g\left( \beta_{1j} | \eta_{1} \right)d\beta_{1j}\int L_{j}\left( \beta_{2j} \right)g\left( \beta_{2j} | \eta_{2} \right)d\beta_{2j} \right)==\prod_{j=1}^{J} \left( \left( 1-\alpha_{10}-\alpha_{01}-\alpha_{11} \right)+\alpha_{10}L_{*j1}\left( \eta_{1} \right)+\alpha_{01}L_{*j2}\left( \eta_{2} \right)+\alpha_{11}L_{*j1}\left( \eta_{1} \right)L_{*j2}\left( \eta_{2} \right) \right)$$

The posterior probability a gene is associated with cancer 1 given the estimates $\alpha_{10}$, $\alpha_{01}$, $\alpha_{11}$, $\eta_{1}$and $\eta_{2}$, is: $P\left( Risk gene for cancer 1 | \mathrm{Data} \right)=\frac{\alpha_{10}L_{1*j}+\alpha_{11}L_{1*j}L_{2*j}}{\left( 1-\alpha_{10}-\alpha_{01}-\alpha_{11} \right)+\alpha_{10}L_{1*j}+\alpha_{01}L_{2*j}+\alpha_{11}L_{1*j}L_{2*j}}$, and similarly for cancer 2. L_1*_ and L_2*_ both depend on $\eta_{1}$and $\eta_{2}$ respectively.

The posterior probability a gene is associated with both cancers is:

$$P\left( Risk gene for both cancers | \mathrm{Data} \right)=\frac{\alpha_{11}L_{1*j}L_{2*j}}{\left( 1-\alpha_{10}-\alpha_{01}-\alpha_{11} \right)+\alpha_{10}L_{1*j}+\alpha_{01}L_{2*j}+\alpha_{11}L_{1*j}L_{2*j}}$$

We can calculate a likelihood ratio test to test for enrichment of susceptibility to cancer 2 given cancer 1 (or vice versa), by comparing the joint likelihood above to the joint likelihood when the cancers are independent.

|  | | $\boldsymbol{Cancer 1,}\boldsymbol{\beta}_{\boldsymbol{1}\boldsymbol{j}}$ | |
| --- | --- | --- | --- |
|  |  | **0 (not associated)**  $w.p$ $\boldsymbol{(1-}\boldsymbol{\alpha}_{\boldsymbol{1}}\boldsymbol{)}$ | $\boldsymbol{g(}\boldsymbol{\beta}_{\boldsymbol{1}}\boldsymbol{\vert}\boldsymbol{\eta}_{\boldsymbol{1}}\boldsymbol{)}$ **(associated)**  $w.p$ $\boldsymbol{\alpha}_{\boldsymbol{1}}$ |
| $Cancer 2, \beta_{2j}$ | **0 (not associated)**  $w.p$ $\boldsymbol{(1-}\boldsymbol{\alpha}_{\boldsymbol{2}}\boldsymbol{)}$ | (0,0)  $w.p$ $\left( 1-\alpha_{10}-\alpha_{01}{-\alpha}_{11} \right)=(1-\alpha_{1}) (1-\alpha_{2})$ | ($g(\beta_{1}\vert\eta_{1}),0)$  $w.p$ $\alpha_{10}$=$\alpha_{1}(1-\alpha_{2})$ |
|  | $\boldsymbol{g(}\boldsymbol{\beta}_{\boldsymbol{2}}\boldsymbol{\vert}\boldsymbol{\eta}_{\boldsymbol{2}}\boldsymbol{)}$ **(associated)**  $w.p \boldsymbol{\alpha}_{\boldsymbol{2}}$ | (0, $g(\beta_{2}\vert\eta_{2}))$  $w.p \alpha_{01}=\alpha_{2}(1-\alpha_{1})$ | ($g(\beta_{1}\vert\eta_{1}),g(\beta_{2}\vert\eta_{2})$)  $w.p \alpha_{11}$=$\alpha_{1}\alpha_{2}$ |

The odds ratio $\frac{\alpha_{11}(1-\alpha_{10}-\alpha_{01}-\alpha_{11})}{\alpha_{10}\alpha_{01}}=\frac{\alpha_{1}\alpha_{2}(1-\alpha_{1}) (1-\alpha_{2})}{\alpha_{1}\alpha_{2}(1-\alpha_{1}) (1-\alpha_{2})}=1$, and $\alpha_{11}$=$\alpha_{1}\alpha_{2}.$Therefore:

$$L\left( \alpha_{10},\alpha_{01},\alpha_{11},\eta_{1},\eta_{2} \right)=\prod_{j=1}^{J} \left( \left( \frac{\alpha_{01}\alpha_{10}}{\alpha_{11}} \right)+\alpha_{10}L_{*j1}\left( \eta_{1} \right)+\alpha_{01}L_{*j2}\left( \eta_{2} \right)+\alpha_{11}L_{*j1}\left( \eta_{1} \right)L_{*j2}\left( \eta_{2} \right) \right)=\prod_{j=1}^{J} \frac{1}{\alpha_{11}}\left( \left( \alpha_{01}\alpha_{10} \right)+{\alpha_{11}\alpha}_{10}L_{*j1}\left( \eta_{1} \right)+{\alpha_{11}\alpha}_{01}L_{*j2}\left( \eta_{2} \right)+{\alpha_{11}}^{2}L_{*j1}\left( \eta_{1} \right)L_{*j2}\left( \eta_{2} \right) \right)=\prod_{j=1}^{J} \frac{1}{\alpha_{11}}\left( \alpha_{01}+{\alpha_{11}L}_{*j1}\left( \eta_{1} \right) \right)\left( \alpha_{10}+{\alpha_{11}L}_{*j2}\left( \eta_{2} \right) \right)=\prod_{j=1}^{J} \frac{1}{\alpha_{1}\alpha_{2}}\left( \alpha_{2}\left( 1-\alpha_{1} \right)+{\alpha_{1}\alpha_{2}L}_{*j1}\left( \eta_{1} \right) \right)\left( \alpha_{1}\left( 1-\alpha_{2} \right)+{\alpha_{1}\alpha_{2}L}_{*j2}\left( \eta_{2} \right) \right)==\prod_{j=1}^{J} (1-\alpha_{1}+\alpha_{1}L_{*j1}\left( \eta_{1} \right))(1-\alpha_{2}+\alpha_{2}L_{*j1}\left( \eta_{2} \right))$$

i.e., the likelihood simplifies to the product of the individual cancer likelihoods, assuming consistent $\eta_{1}$and $\eta_{2}$. Therefore, the likelihood ratio test is a comparison of the joint log-likelihood to the sum of the log-likelihoods of the separate models, using the chi-square distribution with 1 degree of freedom (df).

##### Joint cancer model, method 2

For some cancer pairs method 1 is difficult to optimise and we therefore consider an alternative method where we fix the marginal parameters $\alpha_{1}$ and $\alpha_{2}$ and estimate $\alpha_{3}=P\left( C_{2} \right|C_{1}), \eta_{1}$ and $\eta_{2}$. We then calculate $\alpha_{01},\alpha_{10},\alpha_{11}$ and $\alpha_{00}$:

$$\alpha_{11}=P\left( C_{1}\cap C_{2} \right)=P\left( C_{2} \right|C_{1})P\left( C_{1} \right)={P\left( C_{1} \right|C_{2})P\left( C_{2} \right)=\alpha}_{1}\alpha_{3}$$

$$\alpha_{01}=P\left( {\sim C}_{1}\cap C_{2} \right)=P\left( C_{2} \right|\sim C_{1})P\left( {\sim C}_{1} \right)={P\left( \sim C_{1} \right|C_{2})P\left( C_{2} \right)=\alpha}_{2}\left( 1-\frac{\alpha_{1}\alpha_{3}}{\alpha_{2}} \right)=\alpha_{2}-\alpha_{1}\alpha_{3}$$

$$\alpha_{10}=P\left( C_{1}\cap{\sim C}_{2} \right)=\alpha_{11}=P\left( \sim C_{2} \right|C_{1})P\left( C_{1} \right)=P\left( C_{1} \right|{\sim C}_{2})P\left( \sim C_{2} \right)=\alpha_{1}(1-\alpha_{3})$$

$$\alpha_{00}=P\left( {\sim C}_{1}\cap{\sim C}_{2} \right)=1-\alpha_{1}-\alpha_{2}+\alpha_{1}\alpha_{3}$$

These values should be similar to the values from joint model method 1 if the values of $\eta_{1}$, $\eta_{2}$,$\alpha_{1}$ and $\alpha_{2}$ are similar.

We set $P\left( C_{2} \right|C_{1})$= $\alpha_{3}$, and can calculate $P\left( C_{1} \right|C_{2})=\frac{\alpha_{1}\alpha_{3}}{\alpha_{2}}$

The equivalent likelihood ratio test is testing $\alpha_{3}=\alpha_{2}$, i.e., $P\left( C_{2} \right|C_{1})=P(C_{2})$.

We note that $\alpha_{1}\alpha_{3}\leq\alpha_{1}$ and $\alpha_{1}\alpha_{3}\leq\alpha_{2}$, i.e., $\alpha_{3}\leq min\left( \frac{\alpha_{2}}{\alpha_{1}}, 1 \right)$.

It therefore makes sense to fit the model with cancer 1 being the cancer with the greatest proportion of genes to be risk associated so that the upper bound of $\alpha_{3}$ can consistently be set at 1. If not, and $\alpha_{3}$ is estimated to be $\geq\frac{\alpha_{2}}{\alpha_{1}}$ then $P\left( C_{1} \right|C_{2})$.

This method was used for bowel-endometrial and lung-ovarian cancer.

##### Maximisation

We maximise each log-likelihood to estimate the parameters using the L-BFGS-B optimisation algorithm in the optim package on R. This is a method by Byrd et. Al, 1995 for large non-linear optimisation problems which allows for multivariate estimation and parameter box constraints, i.e., $0<\alpha<1$^25^.

##### Model extension

The joint model method 1 could be extended to more than 2 cancers e.g., breast, prostate and ovarian cancer which have significant overlap. For 3 cancers we would need to estimate 7 $\alpha$’s and 3 $\eta$’s:

$$L\left( \alpha_{100},\alpha_{010},\alpha_{001},{\alpha_{110},\alpha_{101},\alpha_{011},\alpha_{111},\eta}_{1},\eta_{2},\eta_{3} \right)\propto\prod_{j=1}^{J} \left( \left( 1-\alpha_{100}-\alpha_{010}-\alpha_{001}-\alpha_{110}-\alpha_{101}-\alpha_{011}-\alpha_{111} \right)+\alpha_{100}L_{*j1}\left( \eta_{1} \right)+\alpha_{010}L_{*j2}\left( \eta_{2} \right)+\alpha_{001}L_{*j3}\left( \eta_{3} \right)+\alpha_{110}L_{*j1}\left( \eta_{1} \right)L_{*j2}\left( \eta_{2} \right)+\alpha_{101}L_{*j1}\left( \eta_{1} \right)L_{*j3}\left( \eta_{2} \right)+\alpha_{011}L_{*j2}\left( \eta_{1} \right)L_{*j3}\left( \eta_{2} \right)+\alpha_{111}L_{*j1}\left( \eta_{1} \right)L_{*j2}\left( \eta_{2} \right)L_{*j3}\left( \eta_{2} \right) \right)$$

To account for n cancers, we would need to estimate 2^n^-1 $\alpha$’s and n $\eta$’s.

### Supplementary Tables

#### LRT p-values

**Supplementary Table 8 | A comparison of Wald test and LRT P-values for genes reaching exome-wide significance in the Wald test and with case carriers** $\boldsymbol{\leq}$**5.**

| **Cancer** | **Gene** | **Control** | | **Case** | | **OR (CI)** | **P-value** | |
| --- | --- | --- | --- | --- | --- | --- | --- | --- |
|  |  | **Non-carriers** | **carriers** | **Non-carriers** | **carriers** |  | **Wald** | **LRT** |
| Pancreatic | *MEN1* | 418224 | 2 | 1079 | 2 | 429 (57.4, 3200) | 3.46E-09 | 1.66E-05 |
|  | *RCN2* | 418188 | 38 | 1078 | 3 | 31 (9.53, 101) | 1.17E-08 | 0.000136 |
|  | *YPEL3* | 418215 | 11 | 1079 | 2 | 78 (17.2, 355) | 1.73E-08 | 0.000292 |
|  | *SMC2* | 418177 | 49 | 1078 | 3 | 24.9 (7.74, 80.1) | 7.03E-08 | 0.000258 |
|  | *GNG10* | 418205 | 21 | 1079 | 2 | 39.4 (9.15, 169) | 8.05E-07 | 0.00113 |
|  | *ZNF461* | 418201 | 25 | 1079 | 2 | 32.8 (7.71, 139) | 2.26E-06 | 0.00164 |
| Endometrial | *MLH1* | 225410 | 9 | 1971 | 3 | 40.3 (10.9, 149) | 3.24E-08 | 8.2E-05 |
|  | *ACRV1* | 225388 | 31 | 1970 | 4 | 14.6 (5.14, 41.4) | 4.77E-07 | 0.000225 |
|  | *STK32C* | 225403 | 16 | 1971 | 3 | 21.5 (6.25, 74.2) | 1.15E-06 | 0.000466 |
|  | *PSMC6* | 225417 | 2 | 1972 | 2 | 115 (16, 825) | 2.37E-06 | 0.000239 |
| Ovarian | *IVD* | 225957 | 64 | 1367 | 5 | 12.9 (5.19, 32.2) | 3.84E-08 | 6.26E-05 |
|  | *JAML* | 226000 | 21 | 1369 | 3 | 24 (7.15, 80.7) | 2.75E-07 | 0.000322 |
|  | *KCNAB2* | 226016 | 5 | 1370 | 2 | 68.9 (13.2, 360) | 5.17E-07 | 0.000461 |
|  | *ZFP14* | 226016 | 5 | 1370 | 2 | 64.8 (12.5, 337) | 6.92E-07 | 0.000521 |
|  | *TMEM163* | 226015 | 6 | 1370 | 2 | 54.1 (10.9, 269) | 1.09E-06 | 0.000716 |
|  | *TMEM167A* | 226014 | 7 | 1370 | 2 | 47.2 (9.76, 228) | 1.64E-06 | 0.000916 |
|  | *NHEJ1* | 225993 | 28 | 1369 | 3 | 17.7 (5.36, 58.2) | 2.37E-06 | 0.000782 |
| Oesophagus | *KNL1* | 418238 | 67 | 998 | 4 | 23.7 (8.54, 65.6) | 1.17E-09 | 3.09E-05 |
|  | *IRF2BP2* | 418282 | 23 | 1000 | 2 | 36 (8.31, 156) | 1.66E-06 | 0.00138 |
| Kidney | *FGL2* | 417654 | 12 | 1639 | 2 | 49.3 (10.8, 225) | 4.66E-07 | 0.000768 |
|  | *TTC9* | 417627 | 39 | 1638 | 3 | 20.4 (6.25, 66.5) | 5.78E-07 | 0.000493 |
|  | *EXOC7* | 417536 | 130 | 1636 | 5 | 9.53 (3.88, 23.4) | 8.45E-07 | 0.000251 |
|  | *NCK2* | 417652 | 14 | 1639 | 2 | 39.8 (8.94, 178) | 1.34E-06 | 0.00117 |
|  | *TMEM174* | 417653 | 13 | 1639 | 2 | 39.9 (8.86, 180) | 1.60E-06 | 0.00118 |
| Bladder | *DLX2* | 417873 | 9 | 1423 | 2 | 99.5 (19.9, 498) | 2.13E-08 | 0.00205 |
|  | *ZNF506* | 417873 | 9 | 1423 | 2 | 60.6 (12.4, 296) | 3.98E-07 | 0.000555 |
|  | *CDCP2* | 417714 | 168 | 1420 | 5 | 9.81 (4, 24.1) | 6.25E-07 | 0.000220 |
|  | *TMEM222* | 417836 | 46 | 1422 | 3 | 19.5 (5.95, 63.6) | 9.11E-07 | 0.000572 |
|  | *KDM1A* | 417833 | 49 | 1422 | 3 | 18.3 (5.62, 59.7) | 1.41E-06 | 0.000681 |
|  | *ARHGEF6* | 227079 | 2 | 311 | 1 | 400 (35, 4570) | 1.43E-06 | 0.00203 |
|  | *HR* | 417711 | 171 | 1420 | 5 | 9.01 (3.68, 22.1) | 1.53E-06 | 0.000326 |
|  | *NLRP10* | 417879 | 3 | 1424 | 1 | 293 (28.9, 2970) | 1.54E-06 | 0.00269 |
| Malignant Melanoma | *MED9* | 415117 | 11 | 4175 | 4 | 36.1 (11.5, 114) | 8.62E-10 | 9.20E-06 |
|  | *MRPL44* | 415083 | 45 | 4174 | 5 | 11.6 (4.59, 29.3) | 2.17E-07 | 0.000113 |
|  | *CDKN2A* | 415115 | 13 | 4176 | 3 | 23 (6.53, 80.8) | 1.04E-06 | 0.000413 |
|  | *KLHL32* | 415058 | 70 | 4174 | 5 | 7.14 (2.88, 17.7) | 2.24E-05 | 0.00102 |

#### Single cancer heritability results

**Supplementary Table 9 | Heritability results for breast cancer, including the posterior probability of being disease associated, posterior mean effect sizes and the proportion of the familial relative risk (FRR) explained.** Genes listed have posterior probability >0.1. This analysis uses females and males, incorporating family history data, and adjusts for CNV frequency. Results are sorted by descending posterior probability.

| ***Gene*** | **Posterior probability** | **Posterior mean β** | **Posterior mean e^β^** | **λ** | **%FRR** |
| --- | --- | --- | --- | --- | --- |
| ***BRCA1*** | 1.000 | 2.10 | 8.19 | 1.02084 | 2.976 |
| ***BRCA2*** | 1.000 | 1.70 | 5.51 | 1.02525 | 3.598 |
| ***CHEK2*** | 1.000 | 0.85 | 2.35 | 1.00518 | 0.746 |
| ***PALB2*** | 1.000 | 1.34 | 3.81 | 1.00632 | 0.910 |
| ***ATM*** | 1.000 | 0.78 | 2.18 | 1.00176 | 0.254 |
| ***MAP3K1*** | 0.983 | 1.45 | 4.44 | 1.00076 | 0.109 |
| ***BAP1*** | 0.280 | 1.22 | 3.61 | 1.00011 | 0.015 |
| ***PCDHGB3*** | 0.272 | 0.37 | 1.46 | 1.00007 | 0.010 |
| ***COL12A1*** | 0.188 | 0.76 | 2.20 | 1.00005 | 0.008 |
| ***RNF112*** | 0.177 | 0.84 | 2.41 | 1.00005 | 0.007 |
| ***BARD1*** | 0.144 | 0.65 | 1.96 | 1.00007 | 0.010 |
| ***KLK4*** | 0.118 | 0.66 | 1.97 | 1.00003 | 0.004 |
| ***CYBC1*** | 0.116 | 1.92 | 8.22 | 1.00010 | 0.014 |
|  |  |  | **All genes** | **1.06300** | **8.821** |

**Supplementary Table 10 | Heritability results for bowel cancer, including the posterior probability of being disease associated, posterior mean effect sizes and the proportion of the familial relative risk (FRR) explained.** Genes listed have posterior probability >0.1. This analysis uses females and males, incorporating family history data, and adjusts for CNV frequency. Results are sorted by descending posterior probability.

| **Gene** | **Posterior probability** | **Posterior mean β** | **Posterior mean e^β^** | **λ** | **%FRR** |
| --- | --- | --- | --- | --- | --- |
| ***MSH2*** | 1.000 | 1.89 | 6.71 | 1.00614 | 0.884 |
| ***MSH6*** | 1.000 | 1.71 | 5.57 | 1.00769 | 1.105 |
| ***MLH1*** | 1.000 | 2.86 | 17.56 | 1.02583 | 3.678 |
| ***APC*** | 1.000 | 2.39 | 11.27 | 1.00465 | 0.670 |
| ***GAPDH*** | 0.523 | 1.18 | 3.38 | 1.00034 | 0.049 |
| ***FLCN*** | 0.278 | 0.99 | 2.77 | 1.00021 | 0.030 |
|  |  |  | **All genes** | 1.04750 | 6.697 |

**Supplementary Table 11 | Heritability results for prostate cancer, including the posterior probability of being disease associated, posterior mean effect sizes and the proportion of the familial relative risk (FRR) explained.** Genes listed have posterior probability >0.1. This analysis uses females and males, incorporating family history data, and adjusts for CNV frequency. Results are sorted by descending posterior probability.

| ***Gene*** | **Posterior probability** | **Posterior mean β** | **Posterior mean e^β^** | **λ** | **%FRR** |
| --- | --- | --- | --- | --- | --- |
| ***BRCA2*** | 1.000 | 0.72 | 2.05 | 1.00166 | 0.2399 |
| ***CHEK2*** | 1.000 | 0.44 | 1.56 | 1.00100 | 0.1441 |
| ***ATM*** | 0.999 | 0.54 | 1.73 | 1.00067 | 0.0970 |
| ***PPP5C*** | 0.388 | 0.46 | 1.61 | 1.00008 | 0.0109 |
| ***INVS*** | 0.338 | 0.39 | 1.50 | 1.00006 | 0.0084 |
| ***BET1*** | 0.272 | 0.44 | 1.58 | 1.00004 | 0.0060 |
| ***PNLDC1*** | 0.265 | 0.42 | 1.54 | 1.00004 | 0.0061 |
| ***PPEF2*** | 0.247 | 0.25 | 1.30 | 1.00004 | 0.0055 |
| ***CHID1*** | 0.221 | 0.37 | 1.47 | 1.00003 | 0.0041 |
| ***MICB*** | 0.219 | 0.32 | 1.39 | 1.00003 | 0.0041 |
| ***MYH7*** | 0.217 | 0.42 | 1.55 | 1.00003 | 0.0043 |
| ***TRMT44*** | 0.194 | 0.26 | 1.30 | 1.00003 | 0.0048 |
| ***FSIP2*** | 0.184 | 0.19 | 1.22 | 1.00004 | 0.0051 |
| ***SNTG2*** | 0.161 | 0.29 | 1.36 | 1.00002 | 0.0028 |
| ***TMC2*** | 0.154 | 0.34 | 1.42 | 1.00003 | 0.0050 |
| ***WDR59*** | 0.153 | 0.33 | 1.41 | 1.00003 | 0.0045 |
| ***LYST*** | 0.152 | 0.38 | 1.50 | 1.00002 | 0.0027 |
| ***GEMIN2*** | 0.148 | 0.66 | 2.11 | 1.00001 | 0.0019 |
| ***MFSD8*** | 0.135 | 0.49 | 1.71 | 1.00003 | 0.0041 |
| ***C9orf50*** | 0.128 | 0.53 | 1.81 | 1.00001 | 0.0013 |
| ***NAA11*** | 0.126 | 0.26 | 1.32 | 1.00001 | 0.0018 |
| ***CCDC188*** | 0.121 | 0.29 | 1.35 | 1.00001 | 0.0018 |
| ***OAT*** | 0.119 | 0.32 | 1.40 | 1.00001 | 0.0019 |
| ***BSCL2*** | 0.119 | 0.48 | 1.68 | 1.00001 | 0.0014 |
| ***BEND5*** | 0.118 | 0.39 | 1.52 | 1.00001 | 0.0013 |
| ***PABPN1*** | 0.117 | 0.34 | 1.44 | 1.00001 | 0.0014 |
| ***MAP3K19*** | 0.115 | 0.26 | 1.31 | 1.00001 | 0.0017 |
| ***FOXR1*** | 0.111 | 0.38 | 1.51 | 1.00001 | 0.0010 |
| ***SSNA1*** | 0.111 | 0.29 | 1.36 | 1.00001 | 0.0014 |
| ***FANCM*** | 0.107 | 0.17 | 1.19 | 1.00001 | 0.0018 |
| ***OSGIN1*** | 0.107 | 0.56 | 1.90 | 1.00001 | 0.0014 |
| ***CNPY2*** | 0.106 | 0.39 | 1.53 | 1.00001 | 0.0012 |
| ***SORD*** | 0.105 | 0.20 | 1.22 | 1.00001 | 0.0016 |
| ***DPH1*** | 0.104 | 0.27 | 1.32 | 1.00001 | 0.0013 |
| ***DEPDC4*** | 0.103 | 0.23 | 1.27 | 1.00001 | 0.0014 |
| ***SPG7*** | 0.103 | 0.19 | 1.22 | 1.00001 | 0.0015 |
| ***ATP8B4*** | 0.101 | 0.26 | 1.30 | 1.00001 | 0.0019 |
| ***ADAM15*** | 0.101 | 0.27 | 1.33 | 1.00001 | 0.0014 |
|  |  |  | **All genes** | 1.00751 | 1.0798 |

**Supplementary Table 12 | Heritability results for lung cancer, including the posterior probability of being disease associated, posterior mean effect sizes and the proportion of the familial relative risk (FRR) explained.** Genes listed have posterior probability >0.1. This analysis uses females and males, incorporating family history data, and adjusts for CNV frequency. Results are sorted by descending posterior probability.

| **Gene** | **Posterior probability** | **Posterior mean β** | **Posterior mean e^β^** | **λ** | **%FRR** |
| --- | --- | --- | --- | --- | --- |
| ***ATM*** | 0.187 | 0.36 | 1.44 | 1.000055 | 0.00799 |
|  |  |  | **All genes** | 1.000649 | 0.09354 |

**Supplementary Table 13 | Heritability results for pancreatic cancer, including the posterior probability of being disease associated, posterior mean effect sizes and the proportion of the familial relative risk (FRR) explained.** Genes listed have posterior probability >0.1. This analysis uses females and males and adjusts for CNV frequency. Results are sorted by descending posterior probability.

| **Gene** | **Posterior probability** | **Posterior mean β** | **Posterior mean e^β^** | **λ** | **%FRR** |
| --- | --- | --- | --- | --- | --- |
| ***ATM*** | 1.000 | 1.608 | 5.188 | 1.024699086 | 3.520 |
| ***SEC14L3*** | 0.491 | 1.292 | 4.073 | 1.003474889 | 0.500 |
| ***LTV1*** | 0.377 | 0.915 | 2.662 | 1.001748643 | 0.252 |
| ***MROH6*** | 0.239 | 0.887 | 2.631 | 1.000861016 | 0.124 |
| ***MAN2A2*** | 0.152 | 0.607 | 1.916 | 1.000408525 | 0.059 |
| ***ALDH1L1*** | 0.134 | 0.924 | 2.878 | 1.000642404 | 0.093 |
| ***BRCA2*** | 0.127 | 0.653 | 2.035 | 1.000302181 | 0.044 |
| ***PCNT*** | 0.121 | 0.725 | 2.232 | 1.000320685 | 0.046 |
| ***RCN2*** | 0.117 | 1.264 | 4.838 | 1.000593978 | 0.086 |
| ***PKHD1*** | 0.113 | 0.659 | 2.060 | 1.000256601 | 0.037 |
| ***CYP20A1*** | 0.111 | 0.965 | 3.131 | 1.000596335 | 0.086 |
| ***SMC2*** | 0.109 | 1.202 | 4.499 | 1.000741102 | 0.107 |
| ***CEACAM4*** | 0.108 | 0.950 | 3.072 | 1.000499133 | 0.072 |
| ***SMTNL1*** | 0.107 | 0.819 | 2.550 | 1.000219483 | 0.032 |
|  |  |  | **All genes** | 1.0762 | 32.930 |

**Supplementary Table 14 | Heritability results for oesophagus cancer, including the posterior probability of being disease associated, posterior mean effect sizes and the proportion of the familial relative risk (FRR) explained.** Genes listed have posterior probability >0.1. This analysis uses females and males and adjusts for CNV frequency. Results are sorted by descending posterior probability.

| ***Gene*** | **Posterior probability** | **Posterior mean β** | **Posterior mean e^β^** | **λ** | **%FRR** |
| --- | --- | --- | --- | --- | --- |
| ***NLRP12*** | 0.599 | 0.89 | 2.57 | 1.00310 | 0.446 |
| ***ATM*** | 0.543 | 0.95 | 2.79 | 1.00302 | 0.435 |
| ***ZGRF1*** | 0.267 | 0.65 | 2.03 | 1.00078 | 0.113 |
| ***KNL1*** | 0.213 | 1.19 | 4.35 | 1.00051 | 0.074 |
| ***CAMKMT*** | 0.178 | 0.85 | 2.72 | 1.00053 | 0.077 |
| ***CFTR*** | 0.159 | 0.57 | 1.87 | 1.00031 | 0.045 |
| ***FGF11*** | 0.151 | 0.76 | 2.43 | 1.00024 | 0.035 |
| ***PCDHA8*** | 0.126 | 0.45 | 1.62 | 1.00020 | 0.029 |
| ***VPS13A*** | 0.122 | 0.60 | 1.97 | 1.00018 | 0.026 |
| ***RTN4IP1*** | 0.121 | 0.75 | 2.48 | 1.00016 | 0.024 |
| ***TMPRSS11D*** | 0.120 | 0.59 | 1.96 | 1.00016 | 0.022 |
| ***METTL24*** | 0.114 | 0.58 | 1.92 | 1.00015 | 0.022 |
| ***DTHD1*** | 0.111 | 0.70 | 2.31 | 1.00011 | 0.017 |
| ***DFFA*** | 0.110 | 0.70 | 2.31 | 1.00019 | 0.028 |
| ***CTSF*** | 0.106 | 0.68 | 2.24 | 1.00011 | 0.016 |
| ***DIS3*** | 0.103 | 0.54 | 1.84 | 1.00013 | 0.019 |
|  |  |  | **All genes** | 1.04367 | 6.166 |

**Supplementary Table 15 | Heritability results for endometrial cancer, including the posterior probability of being disease associated, posterior mean effect sizes and the proportion of the familial relative risk (FRR) explained.** Genes listed have posterior probability >0.1. This analysis uses females only and adjusts for CNV frequency. Results are sorted by descending posterior probability.

| **Gene** | **Posterior probability** | **Posterior mean β** | **Posterior mean e^β^** | **λ** | **%FRR** |
| --- | --- | --- | --- | --- | --- |
| ***MSH6*** | 1.000 | 2.94 | 19.01 | 1.1070 | 14.666 |
|  |  |  | **All genes** | 1.1214 | 16.526 |

#### Joint cancer heritability results

**Supplementary Table 17 | Optimisation results for cancer pairs**. This includes the method used (method 1 or method 2 in methods), and optimised values of alpha10=P(C1 n C2'), alpha01=P(C1' n C2), alpha11=P(C1 n C2), eta1 and eta2, as well as the p-value from the likelihood ratio test comparing this model to the model where the cancers are independent.

| **Cancer 1** | **Cancer 2** | **Method** | **alpha10** | **alpha01** | **alpha11** | **eta1** | **eta2** | **LRT** |
| --- | --- | --- | --- | --- | --- | --- | --- | --- |
| **Breast** | **Prostate** | 1 | 0.00000 | 0.00000 | 0.00220 | 1.49 | 2.51 | 1.53E-09 |
| **Breast** | **Ovarian** | 1 | 0.00000 | 0.00000 | 0.00270 | 1.77 | 1.23 | 2.12E-08 |
| **Bowel** | **Endom** | 2 | 0.00020 | 0.00000 | 0.00140 | 1.13 | 0.93 | 3.01E-08 |
| **Breast** | **Pancreas** | 1 | 0.00000 | 0.00000 | 0.00230 | 1.62 | 1.57 | 0.0000230 |
| **Prostate** | **Ovarian** | 1 | 0.00000 | 0.00340 | 0.01700 | 6.04 | 2.06 | 0.000151 |
| **Prostate** | **Pancreas** | 1 | 0.00000 | 0.00000 | 0.00840 | 4.56 | 2.17 | 0.000277 |
| **Breast** | **Lung** | 1 | 0.00013 | 0.00026 | 0.00150 | 1.52 | 6.00 | 0.00144 |
| **Breast** | **Bowel** | 1 | 0.00053 | 0.00027 | 0.00160 | 1.51 | 1.00 | 0.00351 |
| **Pancreas** | **Ovarian** | 1 | 0.00000 | 0.00000 | 0.03000 | 3.09 | 2.25 | 0.00540 |
| **Breast** | **Oesophagus** | 1 | 0.00000 | 0.00980 | 0.00240 | 1.64 | 2.95 | 0.00622 |
| **Lung** | **Pancreas** | 1 | 0.00000 | 0.00000 | 0.00460 | 5.93 | 2.05 | 0.00693 |
| **Bowel** | **Ovarian** | 1 | 0.00000 | 0.00000 | 0.00290 | 1.52 | 1.19 | 0.0102 |
| **Prostate** | **Oesophagus** | 1 | 0.00000 | 0.00000 | 0.01400 | 5.49 | 2.84 | 0.0106 |
| **Pancreas** | **Oesophagus** | 1 | 0.00000 | 0.00000 | 0.00750 | 2.28 | 2.37 | 0.0115 |
| **Lung** | **Prostate** | 1 | 0.00002 | 0.00002 | 0.00250 | 6.00 | 2.01 | 0.0161 |
| **Endom** | **Ovarian** | 1 | 0.00000 | 0.00000 | 0.00390 | 1.51 | 1.28 | 0.0213 |
| **Bowel** | **Pancreas** | 1 | 0.00000 | 0.00000 | 0.00220 | 1.40 | 1.68 | 0.0288 |
| **Lung** | **Oesophagus** | 1 | 0.00000 | 0.00000 | 0.00590 | 5.85 | 2.28 | 0.0339 |
| **Bowel** | **Lung** | 1 | 0.00000 | 0.00000 | 0.00260 | 1.49 | 6.00 | 0.0602 |
| **Bowel** | **Prostate** | 1 | 0.00000 | 0.00000 | 0.00340 | 1.83 | 2.93 | 0.112 |
| **Oesophagus** | **Ovarian** | 1 | 0.00000 | 0.01700 | 0.02600 | 3.36 | 2.51 | 0.311 |
| **Lung** | **Endom** | 1 | 0.00050 | 0.00005 | 0.00200 | 6.00 | 1.50 | 0.399 |
| **Bowel** | **Oesophagus** | 1 | 0.00000 | 0.01300 | 0.00230 | 1.46 | 2.94 | 0.460 |
| **Prostate** | **Endom** | 1 | 0.01300 | 0.00000 | 0.00430 | 5.76 | 1.91 | 0.498 |
| **Lung** | **Ovarian** | 2 | 0.00000 | 0.03350 | 0.00350 | 5.43 | 2.41 | 0.602 |
| **Breast** | **Endom** | 1 | 0.00098 | 0.00180 | 0.00150 | 1.52 | 1.51 | 0.639 |
| **Pancreas** | **Endom** | 1 | 0.00270 | 0.00000 | 0.00150 | 1.52 | 1.51 | 1.00 |
| **Oesophagus** | **Endom** | 1 | 0.02400 | 0.00000 | 0.00150 | 3.12 | 1.27 | 1.00 |

**Supplementary Table 18 |Genes with posterior>0.8 for being associated with both cancer 1 and cancer 2 for at least 1 cancer pair.** The cancer pairs columns are the cancer pairs which had posterior probability >0.8 of the gene being associated with both cancer 1 and cancer 2.

| ***Gene*** | **Cancer pairs** |
| --- | --- |
| ***APC*** | Breast-Bowel, Bowel-Lung, Bowel-Ovarian, Bowel-Pancreas, Bowel-Oesophagus |
| ***ATM*** | Breast-Bowel, Breast-Lung, Breast-Oesophagus, Breast-Ovarian, Breast-Pancreas, Breast-Prostate, Lung-Pancreas, Pancreas-Ovarian, Prostate-Ovarian, Prostate-Pancreas, Bowel-Lung, Bowel-Pancreas, Lung-Oesophagus, Lung-Prostate, Pancreas-Oesophagus, Prostate-Oesophagus, Bowel-Prostate, Oesophagus-Ovarian |
| ***BAP1*** | Breast-Prostate |
| ***BRCA1*** | Breast-Bowel, Breast-Lung, Breast-Oesophagus, Breast-Ovarian, Breast-Pancreas, Breast-Prostate, Pancreas-Ovarian, Prostate-Ovarian, Bowel-Ovarian, Endometrial-Ovarian, Breast-Endometrial |
| ***BRCA2*** | Breast-Lung, Breast-Oesophagus, Breast-Ovarian, Breast-Pancreas, Breast-Prostate, Pancreas-Ovarian, Prostate-Ovarian, Prostate-Pancreas, Bowel-Ovarian, Endometrial-Ovarian, Lung-Prostate, Prostate-Oesophagus, Bowel-Prostate |
| ***CHEK2*** | Breast-Bowel, Breast-Oesophagus, Breast-Ovarian, Breast-Pancreas, Breast-Prostate, Prostate-Ovarian, Prostate-Pancreas, Lung-Prostate, Prostate-Oesophagus, Bowel-Prostate |
| ***MAP3K1*** | Breast-Lung, Breast-Oesophagus, Breast-Ovarian, Breast-Pancreas, Breast-Prostate |
| ***MLH1*** | Bowel-Endometrial, Breast-Bowel, Bowel-Lung, Bowel-Ovarian, Bowel-Pancreas, Bowel-Oesophagus, Bowel-Prostate |
| ***MSH2*** | Bowel-Endometrial, Bowel-Lung, Bowel-Ovarian, Bowel-Pancreas, Bowel-Oesophagus, Bowel-Prostate |
| ***MSH6*** | Bowel-Endometrial, Bowel-Lung, Bowel-Ovarian, Bowel-Pancreas, Endometrial-Ovarian, Bowel-Oesophagus, Bowel-Prostate, Lung-Endometrial, Oesophagus-Endometrial, Pancreas-Endometrial, Prostate-Endometrial |
| ***PALB2*** | Breast-Lung, Breast-Oesophagus, Breast-Ovarian, Breast-Pancreas, Breast-Prostate |

**Supplementary Table 19 | Posterior probabilities for the joint cancer modelling of breast and prostate cancer.** The table includes the posterior probability genes are associated with just breast cancer, just prostate cancer, or both cancers. Results shown are for genes with any posterior probability>0.1 and are sorted by descending posterior probability.

| **Gene** | **P(C1 n C2')** | **P(C1' n C2)** | **P(C1 n C2)** |
| --- | --- | --- | --- |
| ***BRCA2*** | 0.000 | 0.000 | 1.000 |
| ***BRCA1*** | 0.000 | 0.000 | 1.000 |
| ***PALB2*** | 0.000 | 0.000 | 1.000 |
| ***ATM*** | 0.000 | 0.000 | 1.000 |
| ***CHEK2*** | 0.000 | 0.000 | 1.000 |
| ***MAP3K1*** | 0.000 | 0.000 | 0.972 |
| ***BAP1*** | 0.000 | 0.000 | 0.824 |
| ***KLK4*** | 0.000 | 0.000 | 0.320 |
| ***PPP5C*** | 0.000 | 0.000 | 0.293 |
| ***OSGIN1*** | 0.000 | 0.000 | 0.204 |
| ***PNLDC1*** | 0.000 | 0.000 | 0.156 |
| ***GEMIN2*** | 0.000 | 0.000 | 0.154 |
| ***CYBC1*** | 0.000 | 0.000 | 0.124 |
| ***DEGS1*** | 0.000 | 0.000 | 0.113 |
| ***SEC62*** | 0.000 | 0.000 | 0.107 |
| ***SNX2*** | 0.000 | 0.000 | 0.103 |

**Supplementary Table 20 | Posterior probabilities for the joint cancer modelling of breast and ovarian cancer.** The table includes the posterior probability genes are associated with just breast cancer, just ovarian cancer, or both cancers. Results shown are for genes with any posterior probability>0.1 and are sorted by descending posterior probability.

| **Gene** | **P(C1 n C2')** | **P(C1' n C2)** | **P(C1 n C2)** |
| --- | --- | --- | --- |
| ***BRCA2*** | 0.000 | 0.000 | 1.000 |
| ***BRCA1*** | 0.000 | 0.000 | 1.000 |
| ***PALB2*** | 0.000 | 0.000 | 1.000 |
| ***CHEK2*** | 0.000 | 0.000 | 1.000 |
| ***ATM*** | 0.000 | 0.000 | 1.000 |
| ***MAP3K1*** | 0.000 | 0.000 | 0.979 |
| ***BAP1*** | 0.000 | 0.000 | 0.416 |
| ***NHEJ1*** | 0.000 | 0.000 | 0.300 |
| ***RAD51D*** | 0.000 | 0.000 | 0.282 |
| ***BRIP1*** | 0.000 | 0.000 | 0.259 |
| ***SLC35E4*** | 0.000 | 0.000 | 0.241 |
| ***KLK4*** | 0.000 | 0.000 | 0.206 |
| ***VWA2*** | 0.000 | 0.000 | 0.200 |
| ***BARD1*** | 0.000 | 0.000 | 0.139 |
| ***IVD*** | 0.000 | 0.000 | 0.139 |
| ***RNF112*** | 0.000 | 0.000 | 0.133 |
| ***ZNHIT1*** | 0.000 | 0.000 | 0.132 |
| ***COL12A1*** | 0.000 | 0.000 | 0.130 |
| ***PCDHGB3*** | 0.000 | 0.000 | 0.122 |
| ***TGM7*** | 0.000 | 0.000 | 0.117 |
| ***CYBC1*** | 0.000 | 0.000 | 0.113 |
| ***PSRC1*** | 0.000 | 0.000 | 0.110 |
| ***TRMT10B*** | 0.000 | 0.000 | 0.105 |
| ***PLEKHG4*** | 0.000 | 0.000 | 0.104 |

**Supplementary Table 21 | Posterior probabilities for the joint cancer modelling of bowel and endometrial cancer.** The table includes the posterior probability genes are associated with just bowel cancer, just endometrial cancer, or both cancers. Results shown are for genes with any posterior probability>0.1 and are sorted by descending posterior probability.

| **Gene** | **P(C1 n C2')** | **P(C1' n C2)** | **P(C1 n C2)** |
| --- | --- | --- | --- |
| ***MSH6*** | 1.92E-42 | 0 | 1.000 |
| ***MLH1*** | 0.019 | 0.000 | 0.981 |
| ***MSH2*** | 0.020 | 0.000 | 0.980 |
| ***APC*** | 0.481 | 0.000 | 0.519 |
| ***GAPDH*** | 0.475 | 0.000 | 0.182 |
| ***FLCN*** | 0.291 | 0.000 | 0.106 |
| ***RDX*** | 0.012 | 0.000 | 0.128 |
| ***NPNT*** | 0.122 | 0.000 | 0.051 |
| ***ACRV1*** | 0.001 | 0.000 | 0.116 |
| ***MPPE1*** | 0.113 | 0.000 | 0.029 |

**Supplementary Table 22 | Posterior probabilities for the joint cancer modelling of breast and pancreatic cancer.** The table includes the posterior probability genes are associated with just breast cancer, just pancreatic cancer, or both cancers. Results shown are for genes with any posterior probability>0.1 and are sorted by descending posterior probability.

| ***Gene*** | **P(C1 n C2')** | **P(C1' n C2)** | **P(C1 n C2)** |
| --- | --- | --- | --- |
| ***BRCA2*** | 0.000 | 0.000 | 1.000 |
| ***BRCA1*** | 0.000 | 0.000 | 1.000 |
| ***PALB2*** | 0.000 | 0.000 | 1.000 |
| ***ATM*** | 0.000 | 0.000 | 1.000 |
| ***MAP3K1*** | 0.000 | 0.000 | 0.980 |
| ***BAP1*** | 0.000 | 0.000 | 0.393 |
| ***RNF112*** | 0.000 | 0.000 | 0.130 |
| ***PCDHGB3*** | 0.000 | 0.000 | 0.146 |
| ***KLK4*** | 0.000 | 0.000 | 0.192 |
| ***COL12A1*** | 0.000 | 0.000 | 0.131 |
| ***CYBC1*** | 0.000 | 0.000 | 0.122 |
| ***CHEK2*** | 0.000 | 0.000 | 1.000 |
| ***RCN2*** | 0.000 | 0.000 | 0.144 |

**Supplementary Table 23 | Posterior probabilities for the joint cancer modelling of prostate and ovarian cancer.** The table includes the posterior probability genes are associated with just prostate cancer, just ovarian cancer, or both cancers. Results shown are for genes with any posterior probability>0.1 and are sorted by descending posterior probability.

| ***Gene*** | **P(C1 n C2')** | **P(C1' n C2)** | **P(C1 n C2)** |
| --- | --- | --- | --- |
| ***BRCA2*** | 0 | 1.47E-12 | 1.000 |
| ***CHEK2*** | 0 | 3.49E-08 | 1.000 |
| ***ATM*** | 0 | 5.50E-06 | 1.000 |
| ***BRCA1*** | 0 | 0.194 | 0.806 |
| ***IVD*** | 0 | 0.119 | 0.468 |
| ***FKBP6*** | 0 | 0.021 | 0.421 |
| ***ANO2*** | 0 | 0.031 | 0.372 |
| ***PPP5C*** | 0 | 0.002 | 0.297 |
| ***BET1*** | 0 | 0.003 | 0.292 |
| ***SLC35E4*** | 0 | 0.102 | 0.263 |
| ***PLEKHG4*** | 0 | 0.076 | 0.261 |
| ***ESYT1*** | 0 | 0.032 | 0.260 |
| ***REXO5*** | 0 | 0.089 | 0.254 |
| ***NEK11*** | 0 | 0.029 | 0.250 |
| ***DPH1*** | 0 | 0.008 | 0.238 |
| ***SORD*** | 0 | 0.008 | 0.223 |
| ***OIP5*** | 0 | 0.018 | 0.214 |
| ***PNLDC1*** | 0 | 0.002 | 0.203 |
| ***CHID1*** | 0 | 0.003 | 0.201 |
| ***PNLIP*** | 0 | 0.032 | 0.198 |
| ***BRIP1*** | 0 | 0.038 | 0.195 |
| ***INVS*** | 0 | 0.001 | 0.188 |
| ***PALB2*** | 0 | 0.009 | 0.175 |
| ***WDFY4*** | 0 | 0.010 | 0.170 |
| ***JAML*** | 0 | 0.034 | 0.169 |
| ***MSH6*** | 0 | 0.025 | 0.162 |
| ***MICB*** | 0 | 0.002 | 0.161 |
| ***R3HCC1L*** | 0 | 0.026 | 0.161 |
| ***TMC2*** | 0 | 0.004 | 0.161 |
| ***ADH1B*** | 0 | 0.033 | 0.151 |
| ***PPEF2*** | 0 | 0.002 | 0.147 |
| ***WDR59*** | 0 | 0.003 | 0.147 |
| ***CEACAM20*** | 0 | 0.025 | 0.146 |
| ***ELP4*** | 0 | 0.050 | 0.146 |
| ***RELT*** | 0 | 0.022 | 0.146 |
| ***TRMT44*** | 0 | 0.002 | 0.146 |
| ***C9orf50*** | 0 | 0.004 | 0.145 |
| ***UBOX5*** | 0 | 0.022 | 0.144 |
| ***ERCC3*** | 0 | 0.006 | 0.144 |
| ***MYH7*** | 0 | 0.002 | 0.143 |
| ***SSNA1*** | 0 | 0.004 | 0.139 |
| ***DOCK2*** | 0 | 0.019 | 0.136 |
| ***PABPN1*** | 0 | 0.004 | 0.133 |
| ***ITGB3*** | 0 | 0.006 | 0.132 |
| ***TRIT1*** | 0 | 0.015 | 0.130 |
| ***GEMIN2*** | 0 | 0.003 | 0.126 |
| ***TMPRSS6*** | 0 | 0.007 | 0.118 |
| ***POLE*** | 0 | 0.020 | 0.113 |
| ***FSIP2*** | 0 | 0.002 | 0.113 |
| ***BEND5*** | 0 | 0.003 | 0.112 |
| ***NHEJ1*** | 0 | 0.030 | 0.111 |
| ***IFNL1*** | 0 | 0.007 | 0.111 |
| ***IFT57*** | 0 | 0.009 | 0.110 |
| ***PIGV*** | 0 | 0.006 | 0.109 |
| ***BRD1*** | 0 | 0.023 | 0.108 |
| ***LMF2*** | 0 | 0.005 | 0.107 |
| ***MARK3*** | 0 | 0.025 | 0.107 |
| ***SNX2*** | 0 | 0.005 | 0.106 |
| ***SPG7*** | 0 | 0.004 | 0.105 |
| ***MXD3*** | 0 | 0.007 | 0.105 |
| ***ZNHIT1*** | 0 | 0.027 | 0.104 |
| ***MFSD8*** | 0 | 0.003 | 0.103 |
| ***PITPNM1*** | 0 | 0.019 | 0.103 |
| ***RAD51D*** | 0 | 0.028 | 0.101 |
| ***FAM161A*** | 0 | 0.011 | 0.100 |
| ***ZRANB3*** | 0 | 0.028 | 0.100 |

**Supplementary Table 24 | Posterior probabilities for the joint cancer modelling of prostate and pancreatic cancer.** The table includes the posterior probability genes are associated with just prostate cancer, just pancreatic cancer, or both cancers. Results shown are for genes with any posterior probability>0.1 and are sorted by descending posterior probability.

| ***Gene*** | **P(C1 n C2')** | **P(C1' n C2)** | **P(C1 n C2)** |
| --- | --- | --- | --- |
| ***BRCA2*** | 0.000 | 0.000 | 1.000 |
| ***ATM*** | 0.000 | 0.000 | 1.000 |
| ***CHEK2*** | 0.000 | 0.000 | 1.000 |
| ***SEC14L3*** | 0.000 | 0.000 | 0.745 |
| ***INVS*** | 0.000 | 0.000 | 0.481 |
| ***GEMIN2*** | 0.000 | 0.000 | 0.232 |
| ***PPP5C*** | 0.000 | 0.000 | 0.210 |
| ***BET1*** | 0.000 | 0.000 | 0.183 |
| ***C9orf50*** | 0.000 | 0.000 | 0.148 |
| ***PALB2*** | 0.000 | 0.000 | 0.147 |
| ***LYST*** | 0.000 | 0.000 | 0.145 |
| ***PNLDC1*** | 0.000 | 0.000 | 0.128 |
| ***SMC2*** | 0.000 | 0.000 | 0.114 |
| ***PACSIN3*** | 0.000 | 0.000 | 0.112 |
| ***MYH7*** | 0.000 | 0.000 | 0.110 |
| ***RCN2*** | 0.000 | 0.000 | 0.110 |
| ***SAG*** | 0.000 | 0.000 | 0.105 |

**Supplementary Table 25 | Posterior probabilities for the joint cancer modelling of breast and lung cancer.** The table includes the posterior probability genes are associated with just breast cancer, just lung cancer, or both cancers. Results shown are for genes with any posterior probability>0.1 and are sorted by descending posterior probability.

| ***Gene*** | **P(C1 n C2')** | **P(C1' n C2)** | **P(C1 n C2)** |
| --- | --- | --- | --- |
| ***ATM*** | 0.000979 | 2.74E-16 | 0.999 |
| ***BRCA2*** | 0.00443 | 4.25E-150 | 0.996 |
| ***BRCA1*** | 0.0760 | 7.27E-77 | 0.924 |
| ***MAP3K1*** | 0.0734 | 5.77E-06 | 0.906 |
| ***PALB2*** | 0.145 | 1.58E-45 | 0.855 |
| ***CHEK2*** | 0.290 | 9.24E-43 | 0.710 |
| ***BAP1*** | 0.0160 | 0.000272 | 0.270 |
| ***PCDHGB3*** | 0.0141 | 0.000199 | 0.149 |
| ***LZTR1*** | 0.00194 | 0.00108 | 0.104 |

**Supplementary Table 26 | Posterior probabilities for the joint cancer modelling of breast and bowel cancer.** The table includes the posterior probability genes are associated with just breast cancer, just bowel cancer, or both cancers. Results shown are for genes with any posterior probability>0.1 and are sorted by descending posterior probability.

| **Gene** | **P(C1 n C2')** | **P(C1' n C2)** | **P(C1 n C2)** |
| --- | --- | --- | --- |
| ***ATM*** | 0.0158 | 2.62E-16 | 0.984 |
| ***BRCA1*** | 0.0218 | 7.39E-77 | 0.978 |
| ***MLH1*** | 4.84E-17 | 0.149 | 0.851 |
| ***APC*** | 3.67E-11 | 0.183 | 0.817 |
| ***CHEK2*** | 0.198 | 1.01E-42 | 0.802 |
| ***BARD1*** | 0.00672 | 0.00211 | 0.791 |
| ***BRCA2*** | 0.310 | 2.84E-150 | 0.690 |
| ***MSH6*** | 1.74E-31 | 0.346 | 0.654 |
| ***MSH2*** | 3.24E-20 | 0.346 | 0.654 |
| ***MAP3K1*** | 0.453 | 3.18E-06 | 0.516 |
| ***PALB2*** | 0.491 | 9.12E-46 | 0.509 |
| ***GAPDH*** | 0.000227 | 0.0825 | 0.491 |
| ***BAP1*** | 0.0610 | 0.000268 | 0.274 |
| ***NPNT*** | 0.000825 | 0.0162 | 0.188 |
| ***CTTNBP2NL*** | 0.00265 | 0.00390 | 0.134 |

**Supplementary Table 28 | Posterior probabilities for the joint cancer modelling of breast and oesophagus cancer.** The table includes the posterior probability genes are associated with just breast cancer, just oesophagus cancer, or both cancers. Results shown are for genes with any posterior probability>0.1 and are sorted by descending posterior probability.

| **Gene** | **P(C1 n C2')** | **P(C1' n C2)** | **P(C1 n C2)** |
| --- | --- | --- | --- |
| ***BRCA2*** | 0.000 | 1.14E-148 | 1.000 |
| ***BRCA1*** | 0.000 | 2.21E-75 | 1.000 |
| ***PALB2*** | 0.000 | 4.74E-44 | 1.000 |
| ***CHEK2*** | 0.000 | 3.15E-41 | 1.000 |
| ***ATM*** | 0.000 | 6.57E-15 | 1.000 |
| ***MAP3K1*** | 0.000 | 0.000 | 0.982 |
| ***NLRP12*** | 0.000 | 0.399 | 0.043 |
| ***BAP1*** | 0.000 | 0.007 | 0.268 |
| ***RNF112*** | 0.000 | 0.010 | 0.199 |
| ***PCDHGB3*** | 0.000 | 0.005 | 0.171 |
| ***KLK4*** | 0.000 | 0.014 | 0.168 |
| ***COL12A1*** | 0.000 | 0.007 | 0.156 |
| ***KNL1*** | 0.000 | 0.146 | 0.012 |
| ***BARD1*** | 0.000 | 0.009 | 0.146 |
| ***ZGRF1*** | 0.000 | 0.130 | 0.003 |
| ***CYBC1*** | 0.000 | 0.009 | 0.126 |

**Supplementary Table 29 | Posterior probabilities for the joint cancer modelling of lung and pancreatic cancer.** The table includes the posterior probability genes are associated with just lung cancer, just pancreatic cancer, or both cancers. Results shown are for genes with any posterior probability>0.1 and are sorted by descending posterior probability.

| **Gene** | **P(C1 n C2')** | **P(C1' n C2)** | **P(C1 n C2)** |
| --- | --- | --- | --- |
| ***ATM*** | 0.000 | 0.000 | 1.000 |
| ***BRCA2*** | 0.000 | 0.000 | 0.438 |
| ***LTV1*** | 0.000 | 0.000 | 0.163 |
| ***SEC14L3*** | 0.000 | 0.000 | 0.161 |
| ***FANCM*** | 0.000 | 0.000 | 0.106 |

### Supplementary Figures

#### Pancreatic Cancer

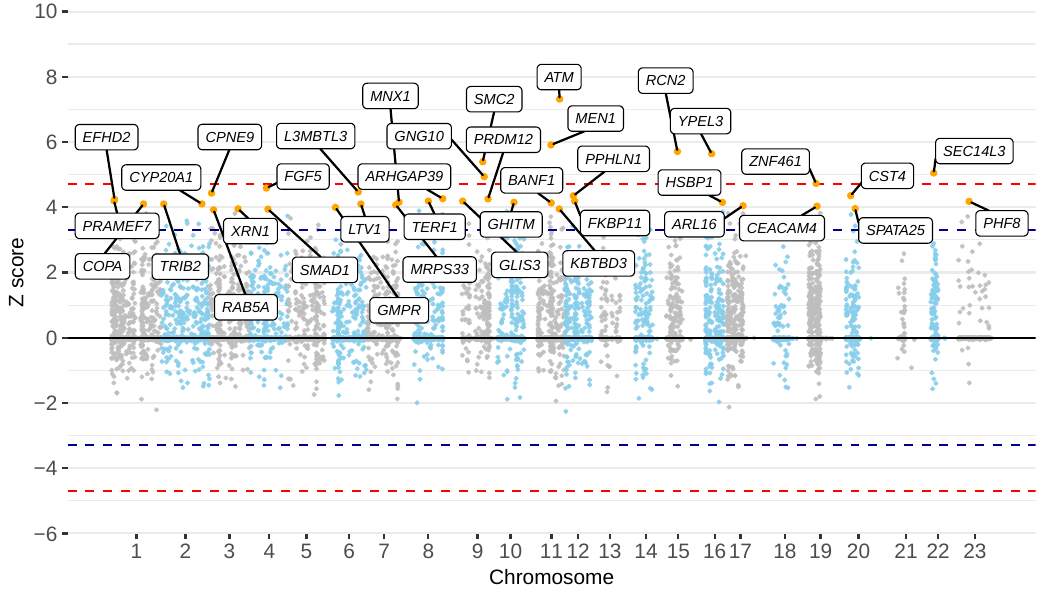

**Figure 1 | Manhattan plot of z scores from assessing the association between protein-truncating variant carriers within genes and pancreatic cancer risk, using model 2.** The x axis is the chromosomal position, and the y axis is the z score from testing H0: β = ln(OR) = 0 (two-tailed) by LRT to the null model. The blue lines correspond to z = ±3.29, P = 0.001, the red lines correspond to z = ±4.71, P = 2.5 × 10−6. All labelled genes are those with P < 0.001. All P-values are unadjusted for multiple testing.

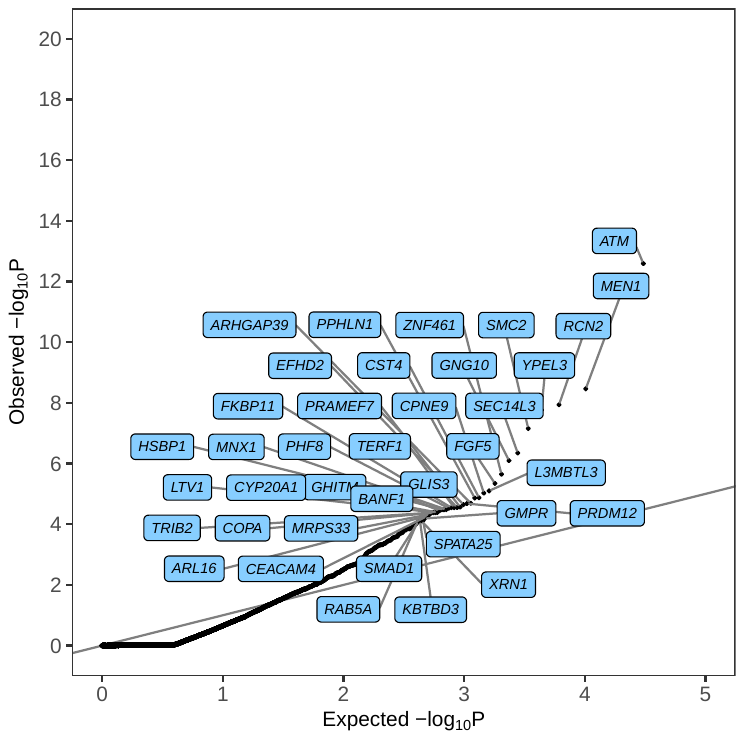

**Figure 2 | Quantile–quantile plot of P values from assessing the association between protein-truncating variant carriers and pancreatic cancer risk.** P-values are from testing H0: β = ln(OR) = 0 by LRT to the null model (two-tailed). The x-axis is the expected log10 P values from the null hypothesis, the y-axis is the observed log10 P value. Highlighted genes have P < 0.0001. Highlighted genes in blue are associated with an increased risk of pancreatic cancer and highlighted genes in cream are associated with decreased risk of pancreatic cancer. All P-values are unadjusted for multiple testing.

#### Endometrial Cancer

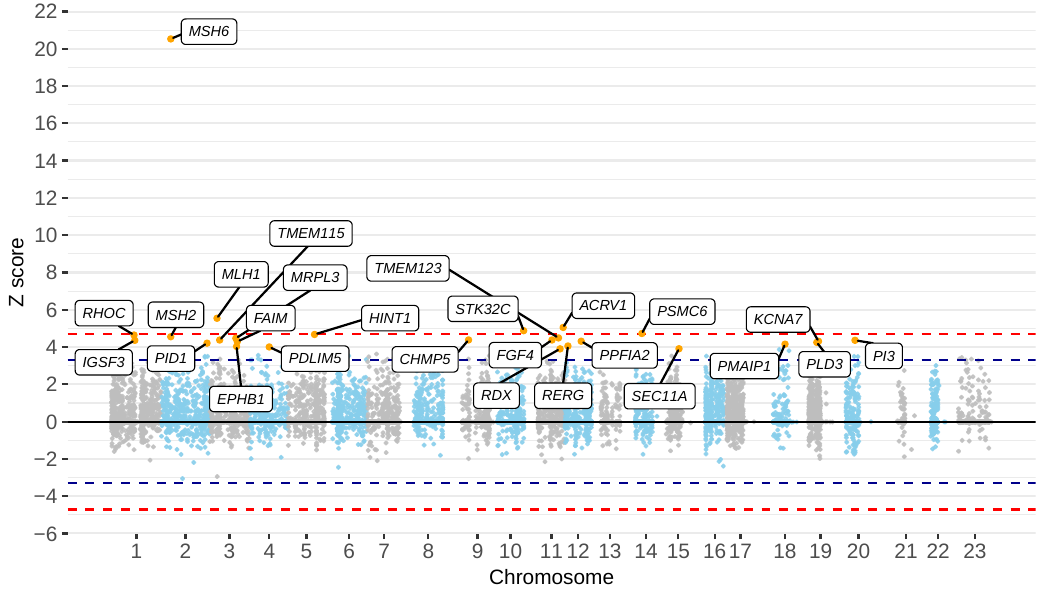

**Figure 3 | Manhattan plot of z scores from assessing the association between protein-truncating variant carriers within genes and endometrial cancer risk, using model 1.** The x axis is the chromosomal position, and the y axis is the z score from testing H0: β = ln(OR) = 0 (two-tailed) by LRT to the null model. The blue lines correspond to z = ±3.29, P = 0.001, the red lines correspond to z = ±4.71, P = 2.5 × 10−6. All labelled genes are those with P < 0.001. All P-values are unadjusted for multiple testing.

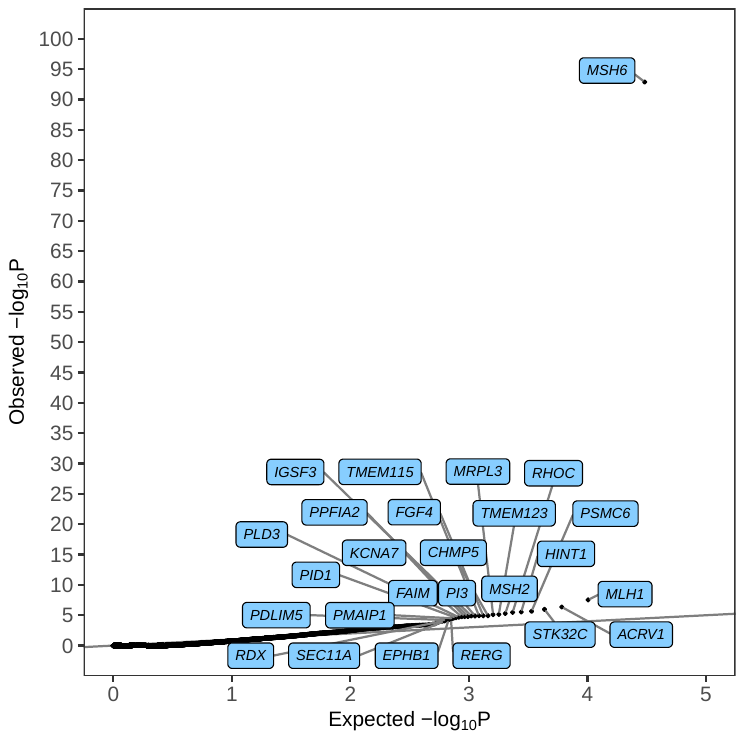

**Figure 4 | Quantile–quantile plot of P values from assessing the association between protein-truncating variant carriers and endometrial cancer risk.** P-values are from testing H0: β = ln(OR) = 0 by LRT to the null model (two-tailed). The x-axis is the expected log10 P values from the null hypothesis, the y-axis is the observed log10 P value. Highlighted genes have P < 0.0001. Highlighted genes in blue are associated with an increased risk of endometrial cancer and highlighted genes in cream are associated with decreased risk of endometrial cancer. All P-values are unadjusted for multiple testing.

#### Ovarian Cancer

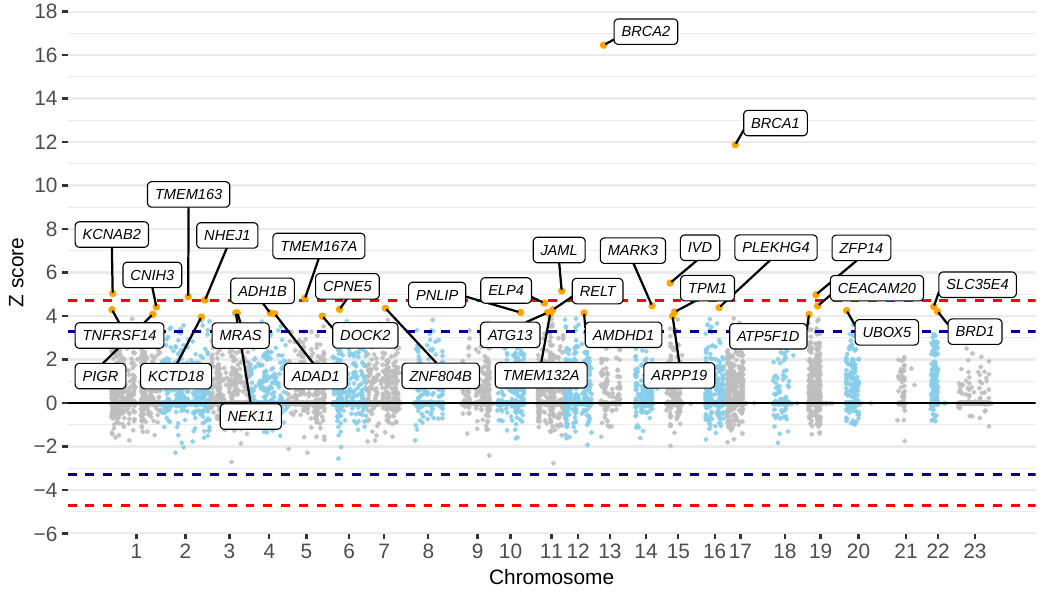

**Figure 5 | Manhattan plot of z scores from assessing the association between protein-truncating variant carriers within genes and ovarian cancer risk, using model 1.** The x axis is the chromosomal position, and the y axis is the z score from testing H0: β = ln(OR) = 0 (two-tailed) by LRT to the null model. The blue lines correspond to z = ±3.29, P = 0.001, the red lines correspond to z = ±4.71, P = 2.5 × 10−6. All labelled genes are those with P < 0.001. All P-values are unadjusted for multiple testing.

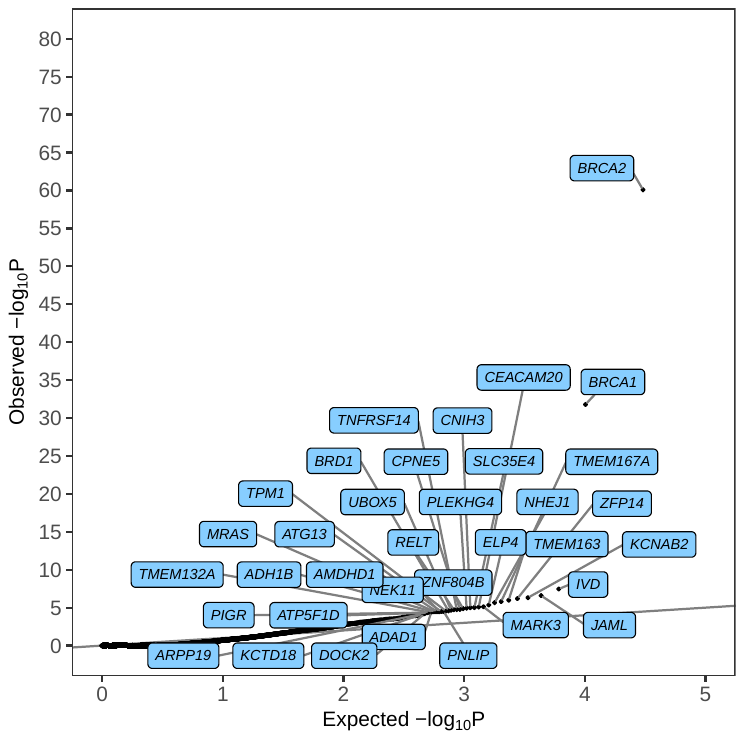

**Figure 6 | Quantile–quantile plot of P values from assessing the association between protein-truncating variant carriers and ovarian cancer risk.** P-values are from testing H0: β = ln(OR) = 0 by LRT to the null model (two-tailed). The x-axis is the expected log10 P values from the null hypothesis, the y-axis is the observed log10 P value. Highlighted genes have P < 0.0001. Highlighted genes in blue are associated with an increased risk of ovarian cancer and highlighted genes in cream are associated with decreased risk of ovarian cancer. All P-values are unadjusted for multiple testing

#### Oesophagus Cancer

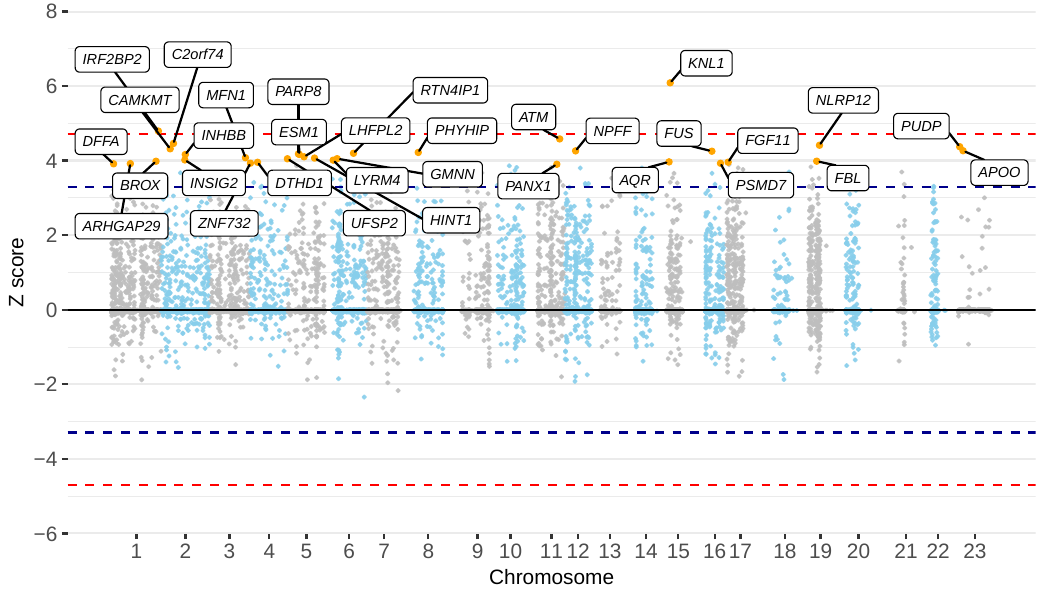

**Figure 7 | Manhattan plot of z scores from assessing the association between protein-truncating variant carriers within genes and oesophagus cancer risk, using model 2.** The x axis is the chromosomal position, and the y axis is the z score from testing H0: β = ln(OR) = 0 (two-tailed) by LRT to the null model. The blue lines correspond to z = ±3.29, P = 0.001, the red lines correspond to z = ±4.71, P = 2.5 × 10−6. All labelled genes are those with P < 0.001. All P-values are unadjusted for multiple testing.

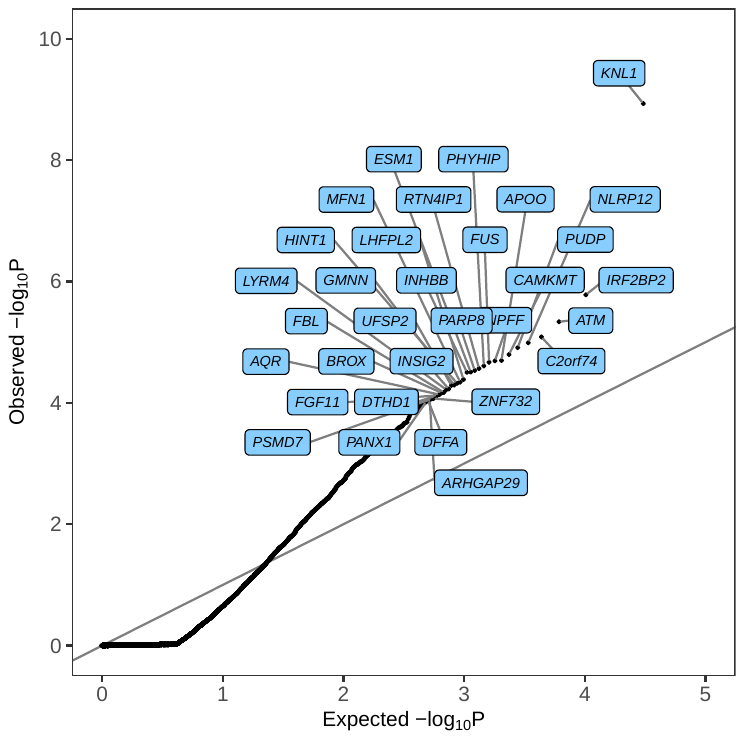

**Figure 8 | Quantile–quantile plot of P values from assessing the association between protein-truncating variant carriers and oesophagus cancer risk.** P-values are from testing H0: β = ln(OR) = 0 by LRT to the null model (two-tailed). The x-axis is the expected log10 P values from the null hypothesis, the y-axis is the observed log10 P value. Highlighted genes have P < 0.0001. Highlighted genes in blue are associated with an increased risk of oesophagus cancer and highlighted genes in cream are associated with decreased risk of oesophagus cancer. All P-values are unadjusted for multiple testing.

#### Kidney Cancer

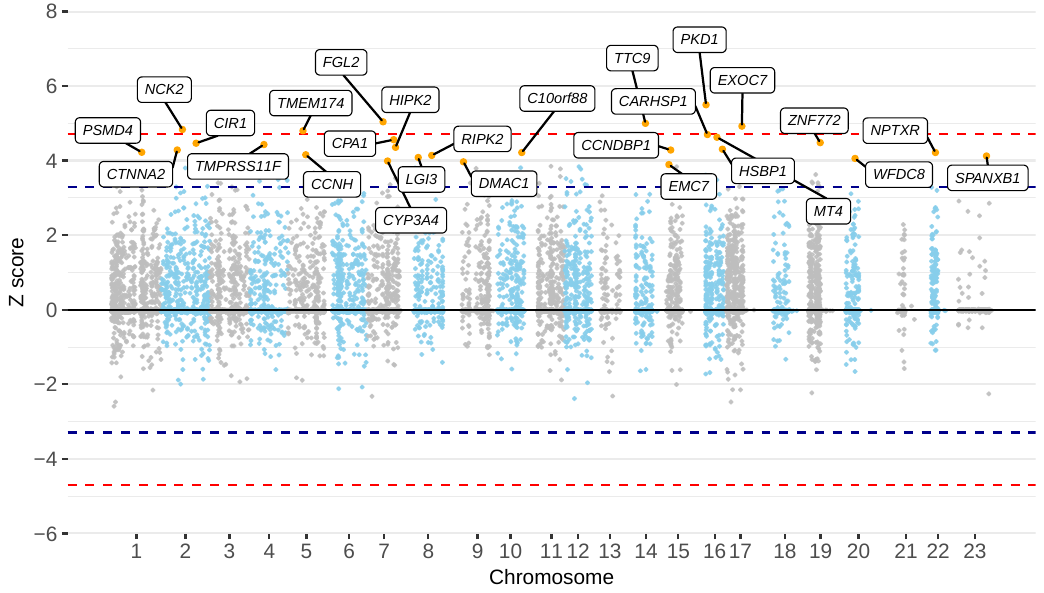

**Figure 9 | Manhattan plot of z scores from assessing the association between protein-truncating variant carriers within genes and kidney cancer risk, using model 2.** The x axis is the chromosomal position, and the y axis is the z score from testing H0: β = ln(OR) = 0 (two-tailed) by LRT to the null model. The blue lines correspond to z = ±3.29, P = 0.001, the red lines correspond to z = ±4.71, P = 2.5 × 10−6. All labelled genes are those with P < 0.001. All P-values are unadjusted for multiple testing.

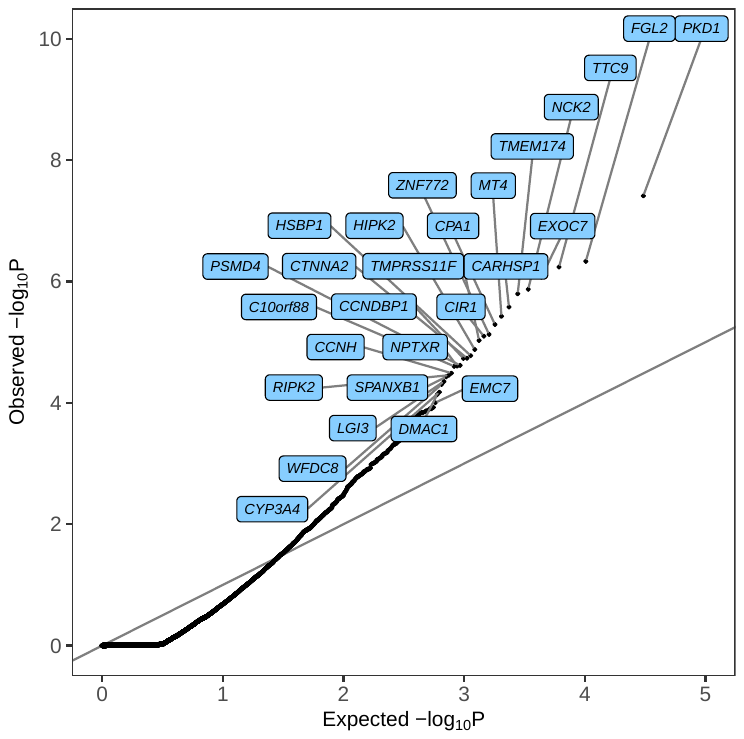

**Figure 10 | Quantile–quantile plot of P values from assessing the association between protein-truncating variant carriers and kidney cancer risk.** P-values are from testing H0: β = ln(OR) = 0 by LRT to the null model (two-tailed). The x-axis is the expected log10 P values from the null hypothesis, the y-axis is the observed log10 P value. Highlighted genes have P < 0.0001. Highlighted genes in blue are associated with an increased risk of kidney cancer and highlighted genes in cream are associated with decreased risk of kidney cancer. All P-values are unadjusted for multiple testing.

#### Bladder Cancer

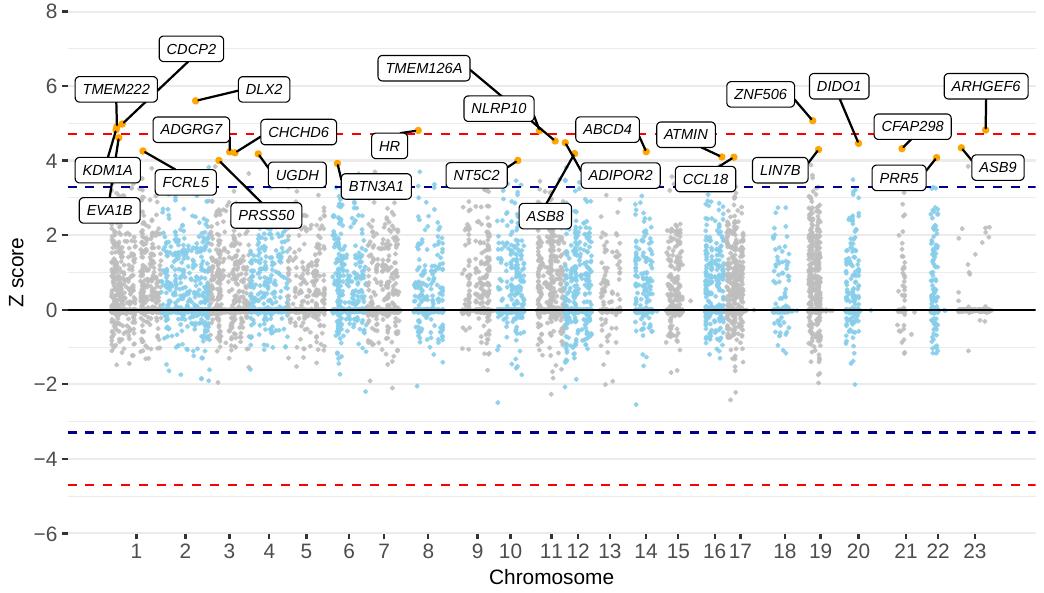

**Figure 11 | Manhattan plot of z scores from assessing the association between protein-truncating variant carriers within genes and bladder cancer risk, using model 2.** The x axis is the chromosomal position, and the y axis is the z score from testing H0: β = ln(OR) = 0 (two-tailed) by LRT to the null model. The blue lines correspond to z = ±3.29, P = 0.001, the red lines correspond to z = ±4.71, P = 2.5 × 10−6. All labelled genes are those with P < 0.001. All P-values are unadjusted for multiple testing.

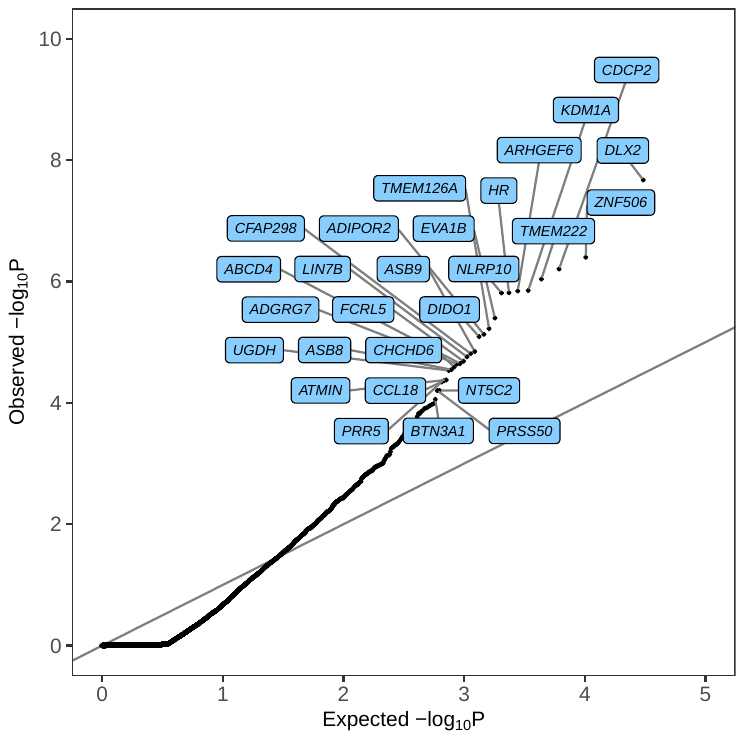

**Figure 12 | Quantile–quantile plot of P values from assessing the association between protein-truncating variant carriers and kidney cancer risk.** P-values are from testing H0: β = ln(OR) = 0 by LRT to the null model (two-tailed). The x-axis is the expected log10 P values from the null hypothesis, the y-axis is the observed log10 P value. Highlighted genes have P < 0.0001. Highlighted genes in blue are associated with an increased risk of kidney cancer and highlighted genes in cream are associated with decreased risk of kidney cancer. All P-values are unadjusted for multiple testing.

#### Malignant Melanoma

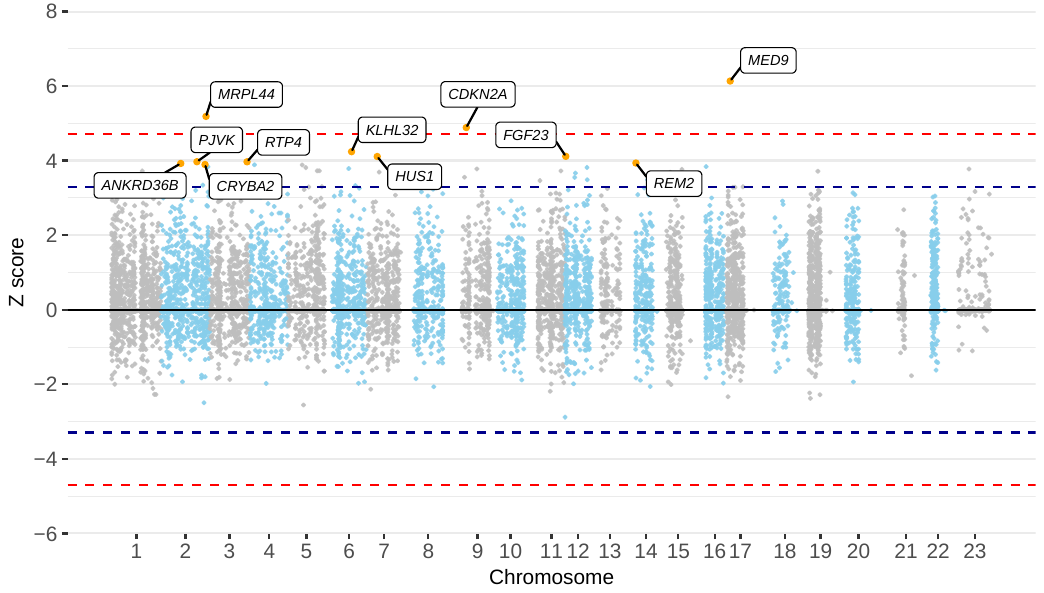

**Figure 13 | Manhattan plot of z scores from assessing the association between protein-truncating variant carriers within genes and malignant melanoma risk, using model 2.** The x axis is the chromosomal position, and the y axis is the z score from testing H0: β = ln(OR) = 0 (two-tailed) by LRT to the null model. The blue lines correspond to z = ±3.29, P = 0.001, the red lines correspond to z = ±4.71, P = 2.5 × 10−6. All labelled genes are those with P < 0.001. All P-values are unadjusted for multiple testing.

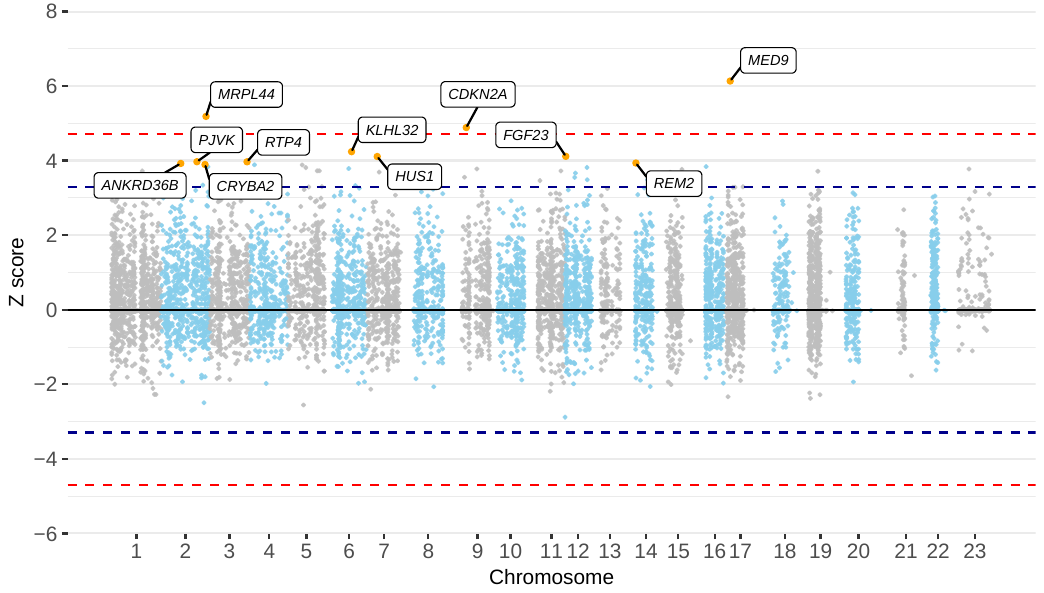

**Figure 14 | Manhattan plot of z-scores from assessing the association between protein-truncating variant carriers within genes and malignant melanoma risk, using model 2.** The x-axis is the chromosomal position, and the y-axis is the z-score from testing H0: β = ln(OR) = 0 (two-tailed) by LRT to the null model. The blue lines correspond to z = ±3.29, P = 0.001, the red lines correspond to z = ±4.71, P = 2.5 × 10−6. All labelled genes are those with P < 0.001. All P-values are unadjusted for multiple testing.

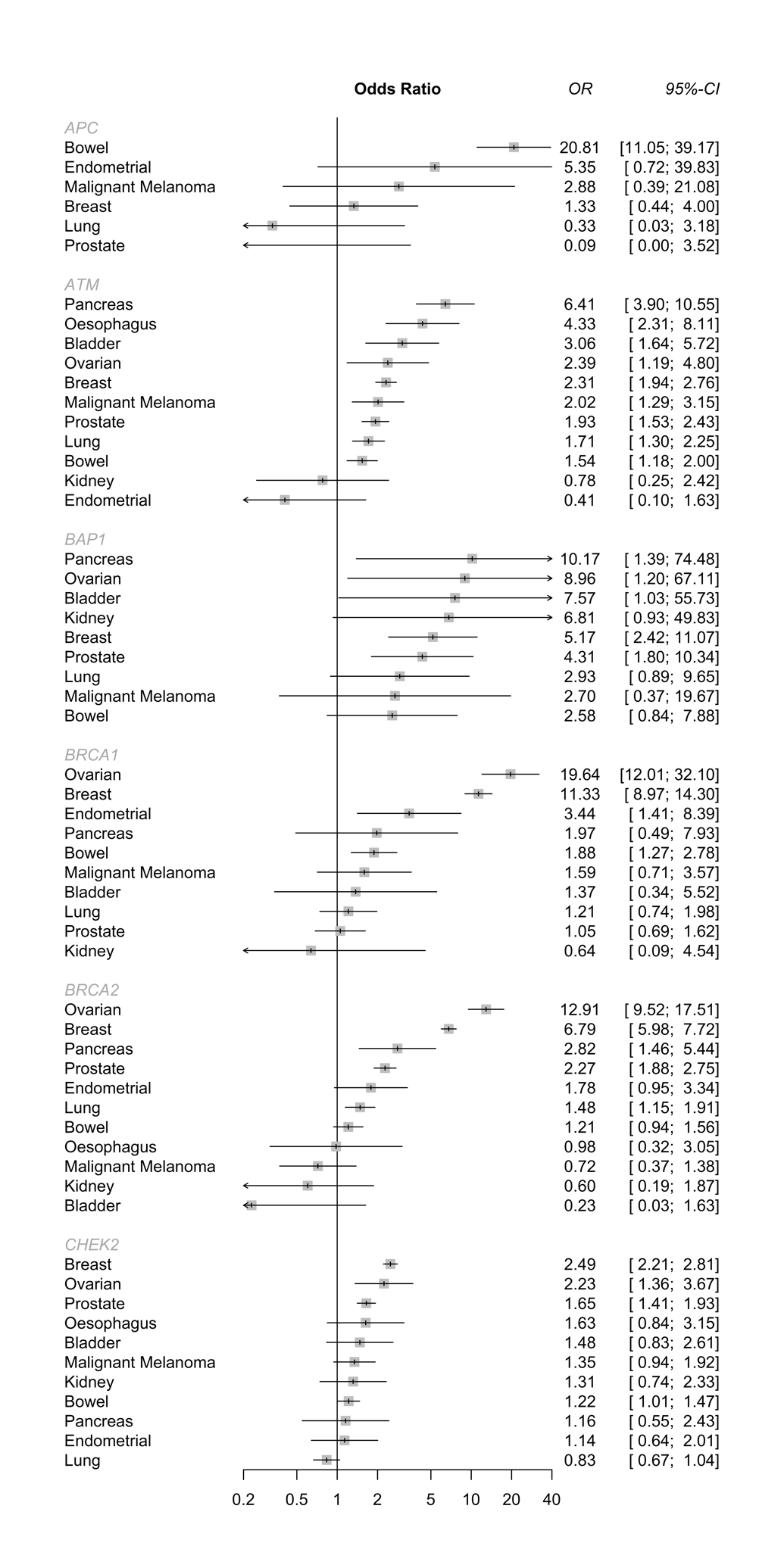

**Figure 15a | Forest plot of PTV burden results across each cancer for the 11 genes with posterior probability > 0.8 in any analysis, and NHEJ1.** For each gene, cancers with 0 case carriers were removed.

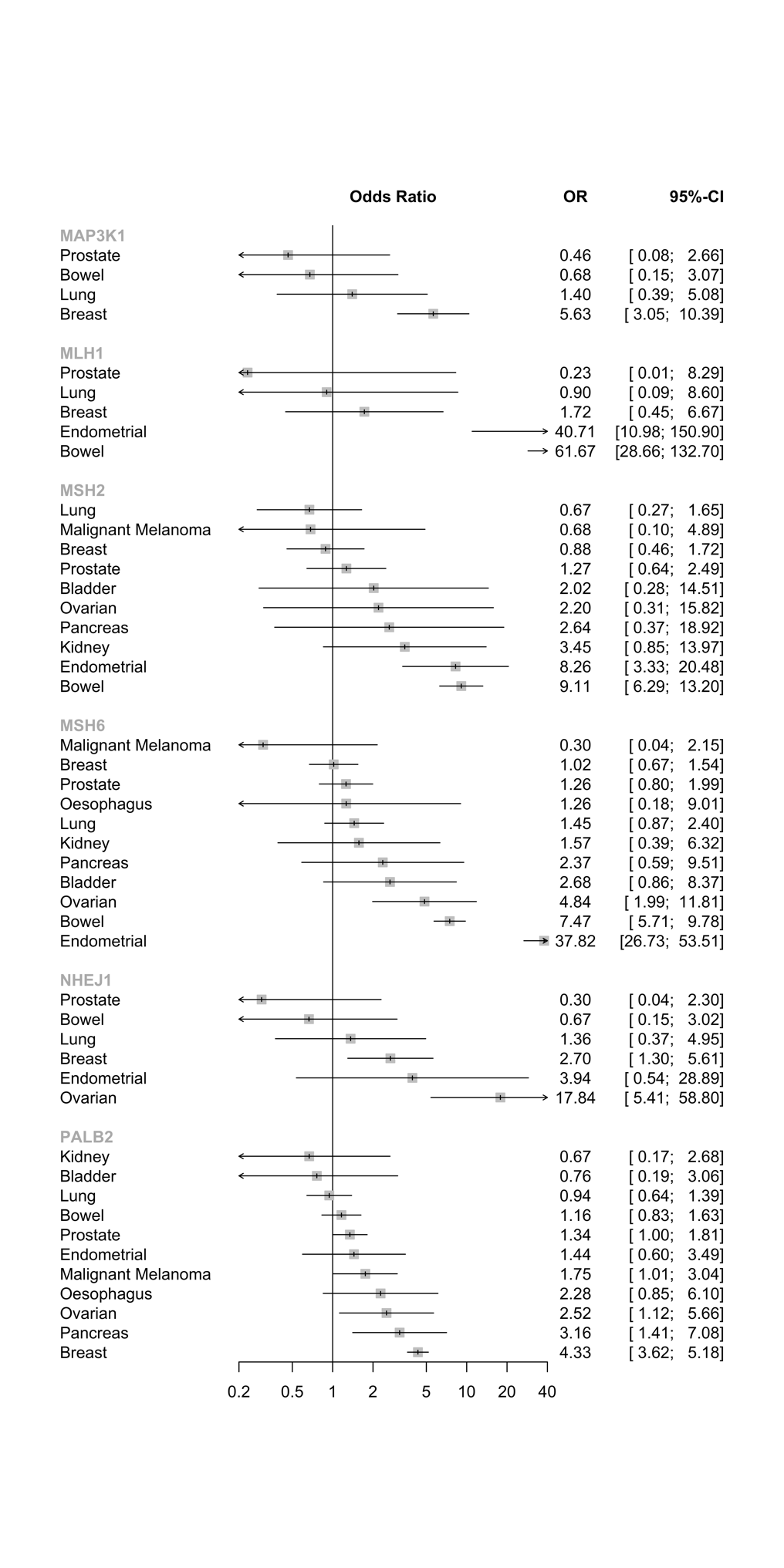

**Figure 15b | Forest plot of PTV burden results across each cancer for the 11 genes with posterior probability > 0.8 in any analysis, and NHEJ1.** For each gene, cancers with 0 case carriers were removed.
